# Comprehensive evaluation of the impact of sociodemographic inequalities on adverse outcomes and excess mortality during the COVID-19 pandemic in Mexico City

**DOI:** 10.1101/2021.03.11.21253402

**Authors:** Neftali Eduardo Antonio-Villa, Luisa Fernandez-Chirino, Julio Pisanty-Alatorre, Javier Mancilla-Galindo, Ashuin Kammar-García, Arsenio Vargas-Vázquez, Armando González-Díaz, Carlos A. Fermín-Martínez, Alejandro Márquez-Salinas, Enrique Cañedo-Guerra, Jessica Paola Bahena-López, Marco Villanueva-Reza, Jessica Márquez-Sánchez, Máximo Ernesto Jaramillo-Molina, Luis Miguel Gutiérrez-Robledo, Omar Yaxmehen Bello-Chavolla

**Affiliations:** Division de Investigación, Instituto Nacional de Geriatría, Mexico City, Mexico; MD/PhD (PECEM), Faculty of Medicine, National Autonomous University of Mexico, Mexico City, Mexico; Faculty of Chemistry, National Autonomous University of Mexico, Mexico City, Mexico; Departamento de Salud Pública, Facultad de Medicina, Universidad Nacional Autónoma de México, Mexico City, Mexico; Instituto Mexicano del Seguro Social, Mexico City, Mexico; Unidad de Investigación UNAM-INC, Instituto Nacional de Cardiología Ignacio Chávez, Mexico City, Mexico; Departamento de Atención Institucional Continua y Urgencias, Instituto Nacional de Ciencias Médicas y Nutrición Salvador Zubirán, Mexico City, Mexico; Sección de Estudios de Posgrado e Investigación, Escuela Superior de Medicina, Instituto Politécnico Nacional, Mexico City, Mexico; Facultad de Ciencias Politicas Sociales y Sociales, National Autonomous University of Mexico, Mexico City, Mexico; Departamento de Infectologia. Instituto Nacional de Ciencias Médicas y Nutrición Salvador Zubirán, Mexico City, Mexico; Departamento de Infectología, Instituto Nacional de Pediatría, Mexico City, Mexico; International Inequalities Institute, London School of Economics, London, UK

**Keywords:** Epidemiology, social inequalities, public health, Mexico, COVID-19, SARS-CoV-2

## Abstract

The impact of the COVID-19 pandemic in Mexico City has been sharp, as several social inequalities coexist with chronic comorbidities. Here, we conducted an in-depth evaluation of the impact of social, municipal, and individual factors on the COVID-19 pandemic in working-age population living in Mexico City. To this end, we used data from the National Epidemiological Surveillance System; furthermore, we used a multidimensional metric, the social lag index (DISLI), to evaluate its interaction with mean urban population density (MUPD) and its impact on COVID-19 rates. Influence DISLI and MUPD on the effect of vehicular mobility policies on COVID-19 rates were also tested. Finally, we assessed the influence of MUPD and DISLI on discrepancies of COVID-19 and non-COVID-19 excess mortality compared with death certificates from the General Civil Registry. We detected vulnerable groups who belonged to economically active sectors and who experienced increased risk of adverse COVID-19 outcomes. The impact of social inequalities transcends individuals and has significant effects at a municipality level, with and interaction between DISLI and MUPD. Marginalized municipalities with high population density experienced an accentuated risk for adverse COVID-19 outcomes. Additionally, policies to reduce vehicular mobility had differential impacts across marginalized municipalities. Finally, we report an under-registry of COVID-19 deaths and significant excess mortality associated with non-COVID-19 deaths closely related to MUPD/DISLI in an ambulatory setting, which could be a negative externality of hospital reconversion. In conclusion, social, individual, and municipality-wide factors played a significant role in shaping the course of the COVID-19 pandemic in Mexico City.

## INTRODUCTION

The COVID-19 pandemic has brought health inequalities worldwide into sharp relief. Although clinical risk factors for severe outcomes and lethality, such as increased age and a high burden of chronic health comorbidities, were identified soon after the first reported cases in Wuhan (1–3), these do not fully explain the disproportionate burden of COVID-19 pandemic experienced by historically marginalized population groups, as has been widely documented in Europe (4–7) and North America (8–14). A possible hypothesis according to the ‘Social Determination of the Health-Disease Process’ (SDHDP) approach expresses that the cumulative and yet dynamic interplay between historically, spatially and socially shaped processes of exposure/imposition, susceptibility and resistance, which dialectically affects health at the singular, particular and general levels, could have contributed within the current world-health situation (15). Going beyond the better-known Social Determinants of Health approach, the SDHDP approach focuses on the social processes that underlie the inequitable distribution of risk factors and poor health (15–17). Similarly, eco-social epidemiology focuses on the cumulative interplay between exposure, susceptibility and resistance which leads to health, and the notion of *embodiment* (18). Both of these approaches, regard empirical data on health disparities along socioeconomic, geospatial, racialized and gendered lines as a starting point to understand the processes that shape population health – the COVID-19 pandemic being no exception.

Mexico City is one of the most economically unequal cities in the continent, with large percentages of the population working in informal conditions and living in poverty, with resulting documented disparities of over 20 years in life expectancy between municipalities in the wider metropolitan region (19). Similarly, a large proportion of the population does not have a proper support network or social security for in-hospital care (20). These factors are further complicated by the high prevalence of cardiometabolic diseases in all age groups in the Mexican population, which have been shown to worsen adverse outcomes attributable to COVID-19 (21–23). Despite the high burden of COVID-19 in Latin America and Mexico in particular, and early predictions of the effect that social inequality would have on the development of the pandemic, research on COVID-19 and social inequality has been comparatively scarce (24,25). We thus aimed to assess the impact of social inequalities on the course of the COVID-19 pandemic in Mexico City. To this end, we explored working occupational roles as an individual determinant on the risk for COVID-19 complications. Next, we evaluated the influence of the municipal social lag as a determinant of epidemiological indicators of the pandemic and the influence of vehicular mobility reduction on them. Finally, we evaluated whether the discrepancies of reported deaths and the excess mortality attributable to both COVID-19 and non-COVID-19 causes was higher for marginalized municipalities within Mexico City.

## METHODS

### Data Sources

#### COVID-19 suspected cases and related outcomes in Mexico City

We analyzed data collected in the National Epidemiological Surveillance Study (NESS). We limited our analyses to the 16 core municipalities in Mexico City. The NESS is an open-source dataset made available by national health authorities which comprises daily-updated suspected COVID-19 cases that have been tested using both real-time RT-PCR according to the Berlin Protocol and rapid antigen testing to confirm SARS-CoV-2 (26), and were certified by the National Institute for Epidemiological Diagnosis and Referral (27,28). COVID-19 suspected case definition and complete data recollection regarding comorbidities, symptoms and sociodemographic variables in the NESS are included in **Supplementary Material**. Dates of symptom onset, hospital admission, and death are available for all cases as well as outpatient or hospitalized management status, diagnosis of clinical pneumonia, ICU admission, and whether the patient required invasive mechanical ventilation (IMV). Asymptomatic cases were defined as subjects who referred no associated symptomatology at admission but had confirmed SARS-CoV-2 infection. Severe outcome was defined as a composite outcome comprising death, requirement for IMV or ICU admission (29).

#### Definitions of working group categories

We accommodated reported occupation within the NESS according to six working groups using definitions by the National Institute of Statistics and Geography (INEGI) as: economically active workers, health-care workers (HCWs), retired workers, home-care related workers, unemployed subjects and students at the point of assessment for suspected SARS-CoV-2 infection. Complete details of the classification and estimated population for each working group are available in **Supplementary Material**. We weighted estimates stratified by working group categories using population of economically active and inactive population >15 years based on trimestral projections from the 2020 National Survey of Occupation and Employment (ENEO) conducted by INEGI (30). Given the increased occupational risk of HCWs for SARS-CoV-2 infection, we used this group as reference to estimate risk for COVID-19 outcomes compared to other occupations (31).

#### Urban Population Density-Independent Social lag estimation

To quantify the impact of socio-demographic inequalities on the course of the COVID-19 pandemic at a municipal level, we used the 2015 social lag index (SLI). The SLI indicates a composite of social lag assessed by the degree of health-care access, economic well-being, and access to basic services within Mexican Municipalities. (16). We also calculated the mean urban population density (MUPD) for the City’s 16 municipalities using the standard method proposed by the National Institute for Geography and Information (INEGI). We observed that SLI was highly correlated to MUPD; since we intended to evaluate inequalities independent or urbanization, we regressed MUPD onto SLI values using linear regression and extracted the residuals to obtain a MUPD-independent SLI index (DISLI)(33). To assess cut-offs for both DISLI and MUPD, we used the Jenks optimization method to identify natural breaks in continuous variables. This allowed us to divide the municipalities into four categories by Low MUPD /Low DISLI, High MUPD /Low DISLI, Low MUPD /High DISLI and High MUPD /High DISLI. Details of DISLI distribution and municipalities classification in Mexico City is presented in **Supplementary Material**. For population-weighted analyses by DISLI categories and for adjustments by MUPD we extracted data from 2020 population projections obtained by the Mexican National Population Council (CONAPO) for Mexico City (34).

#### Vehicular mobility reduction estimates

To determine the heterogeneous impact of social isolation policies on epidemiological rates we used the vehicular mobility data within Mexico City. Vehicular mobility data was acquired from an open-source dataset provided by the Mexico City’s government, which comprises a relative percentual comparison of vehicular mobility with the same day in 2019. These changes represent the vehicular traffic flow captured in Mexico City by transit records according to the Mobility Department (SEMOVI) (35) by week and municipality. Further methodology is presented in **Supplementary Material**. This dataset was last updated on December 31^st^, 2020.

#### Death certificates of COVID-19 and other non-COVID-19 causes

Given that widespread to access to SARS-CoV-2 testing in Mexico City has been limited, a considerable number of infections and infection-related deaths of cases who were likely treated at home have not been consistently recorded (36). Given this underreport and the indirect effects of the restructuring of the Mexican healthcare system, excess mortality is a more reliable metric of the effects of the pandemic in Mexico City. Hence, we estimated excess deaths of suspected COVID-19 and non-COVID-19 causes using an open-source database of monthly updated death certificates captured by the general civil registry (GCR) of Mexico City (37). A complete description of the GCR dataset is presented in **supplementary material**.

### Statistical analysis

#### Population-based statistics

We estimated the incidence, testing, asymptomatic and mortality rates by standardizing cases with their respective population denominators and normalized to rates per 100,000 inhabitants. All rates were plotted for every month since the first reported case on February 28^th^, 2020 to evaluate the progression of the pandemic in Mexico City. A p-value <0.05 was considered as the statistical significance threshold. All analyses were performed using R version 4.0.0.

#### Conditions related to SARS-CoV-2 outcomes in workers in Mexico City

To assess if individual and structural inequalities were associated with hospitalization, IMV, severe outcome and lethality attributable to SARS-CoV-2, we fitted mixed effects Cox Proportional Hazard Risk models, including facility of treatment as a random intercept to account for the variability in case distribution and treatments across municipalities. We excluded all patients who were suspected cases at the time of inclusion without confirmed SARS-CoV-2 infection. Models were selected based on the Bayesian Information Criterion (BIC) and performance was assessed using Harrel’s c-statistic.

#### Mobility trends and its impact in COVID-19 rates across Mexico City

To evaluate the impact of the reduction of vehicular mobility on the COVID-19 pandemic in Mexico City, we used relative mobility indexes according to the corresponding dates of registry and municipality of residence for all confirmed cases. To evaluate whether mobility decrease had any effect on incidence, mortality, severe case, and hospitalization rates according to DISLI quadrants, we fitted multilevel mixed-effect Poisson models adjusted by age, public-policy epidemiologic period, MUPD, and number of comorbidities. Relative mobility was adjusted using restricted cubic splines. These models considered the interaction effects between MUPD/DISLI quadrants with the individually assigned mobility index, with population offset assigned according to sex and corresponding DISLI population as the denominator. To determine if there was temporal causality on the effect of vehicular mobility reduction on COVID-19 rates, Granger causality tests were performed considering relative mobility decrease as both the predictor and as the result. To clarify the nature of the behaviors of these rates in time, and to evaluate their behavior as functions of themselves when adjusted by mobility, we fitted Autorregressive-Distributed Lag (ARDL) models for incidence and mortality rates, dividing data by MUPD/DISLI quadrant and adjusting with work-group categorization. Furthermore, ARDL models were also fitted for both rates but using our worker-group categorization as a classifier with DISLI as a continuous variable adjustment. All ARDL models were validated and best fit according to lag was selected with BIC criteria and Bounds tests were used to evaluate short and long term cointegration. *Estimation of excess mortality of non-COVID-19 deaths*.

To assess the influence of SLI on the underreport of suspected and confirmed COVID-19 deaths in Mexico City, we estimated discrepancies in mortality rates comparing the NESS and GCR death certificates datasets across our MUPD/DISLI categories. To estimate excess mortality, we calculated the difference between the mortality rate of non-COVID-19 deaths by death certificate of 2020 and the average mortality rate for the 2017-2019 period adjusted to the population of each year according to MUPD/DISLI categories. Finally, the impact of MUPD /DISLI categories on underreport and excess mortality was tested using negative binomial regression models, using the population of each MUPD/DISLI categories as offsets to estimate incidence rate ratios (IRR). Models were further adjusted for the dependency and masculinity indexes of non-COVID-19 deaths, which are demographic indicators used to account for the population structure (38).

## RESULTS

### The COVID-19 pandemic in Mexico City

Since the beginning of the pandemic in Mexico City until January 31^st^, 2021 the NESS tested 1,473,746 persons for suspected COVID-19 in 688 medical units, amongst the estimated 9,018,645 (16.3%) inhabitants of Mexico City in 2020. Amongst them, 423,050 (28.7%) had confirmed SARS-CoV-2 infection, of whom 47,478 (3.2%) were asymptomatic cases. The timeline since the first confirmed case in Mexico City along with the phases of government response are presented in **Supplementary Material**. Reported COVID-19 outcomes amongst evaluated cases included 82,771 (5.6%) who required hospitalization, 12,772 who required IMV (0.86%) and 36,732 (2.5%) who had a severe outcome. Case fatality attributable to suspected and confirmed COVID-19 was recorded in 32,076 (2.2%) cases. According to the WHO epidemiological surveillance definition, most evaluated subjects were classified as cases of influenza-like illness (ILI) without SARS-CoV-2 infection (64.9%) and ILI with SARS-CoV-2 infection (24.6%) and to a lesser extent, cases with severe acute respiratory syndrome (SARI) without SARS-CoV-2 infection (6.4%) and SARI with SARS-CoV-2 infection (4.1%). We observed that testing rates have been steadily increasing since April-2020. Consequently, this has led to an increase in the rates of incident and asymptomatic infections. Moreover, we observed peaks in mortality rates in May-2020 and December-2021 (**Supplementary Material**).

### Working group as a social determinant of COVID-19 rates

We found a high proportion of confirmed cases reported as economically active (49.8%), followed by home-related workers (14.7%), other non-specified workers (12%), students (10.7%), HCWs (6.4%), unemployed (3.6%) and retired adults (2.8%). Subjects classified as retired adults, home-related workers and unemployed were older and had increased prevalence of cardiometabolic comorbidities; furthermore, retired adults had an increased proportion of SARS-CoV-2 positive SARI cases at admission. Consistent with previous reports, we observed that HCWs had the highest testing and incidence rates amongst worker categories. Nevertheless, the group of other non-specified workers had the second highest rates in testing, incidence, and the highest rate of asymptomatic cases. Finally, unemployed, other non-specified workers and retired adults similarly had increased mortality rates throughout the pandemic. When analyzing only lethal cases, home-related workers, retired adults, and unemployed subjects were markedly older and had a higher burden of arterial hypertension, diabetes and obesity (**Supplementary Material**).

### Sociodemographic inequalities and their impact in the COVID-19 pandemic

Dividing our study population across MUPD/DISLI categories, we observed that a larger proportion of tested subjects lived in High MUPD/High DISLI (61.8%), followed by Low MUPD/High DISLI (13.9%), High MUPD/Low DISLI (13.5%) and Low MUPD/Low DISLI (10.7%). Subjects living in Low MUPD/Low DISLI municipalities were older and had a higher prevalence of cardiometabolic comorbidities. Moreover, we observed Low MUPD/High DISLI municipalities had the highest proportion SARS-CoV-2 positive SARI cases (**Supplementary Material**). Over the course of the pandemic, we observed a higher age-adjusted testing rate in Low MUPD/High DISLI municipalities; therefore, age-adjusted incidence and asymptomatic rates followed a similar pattern. Nevertheless, over the course of the pandemic, we observed a DISLI-dependent mortality pattern, in which High MUPD/High DISLI and Low MUPD/High DISLI municipalities, had the highest age-adjusted mortality rates (**Figure 1**).

**Figure 1:**
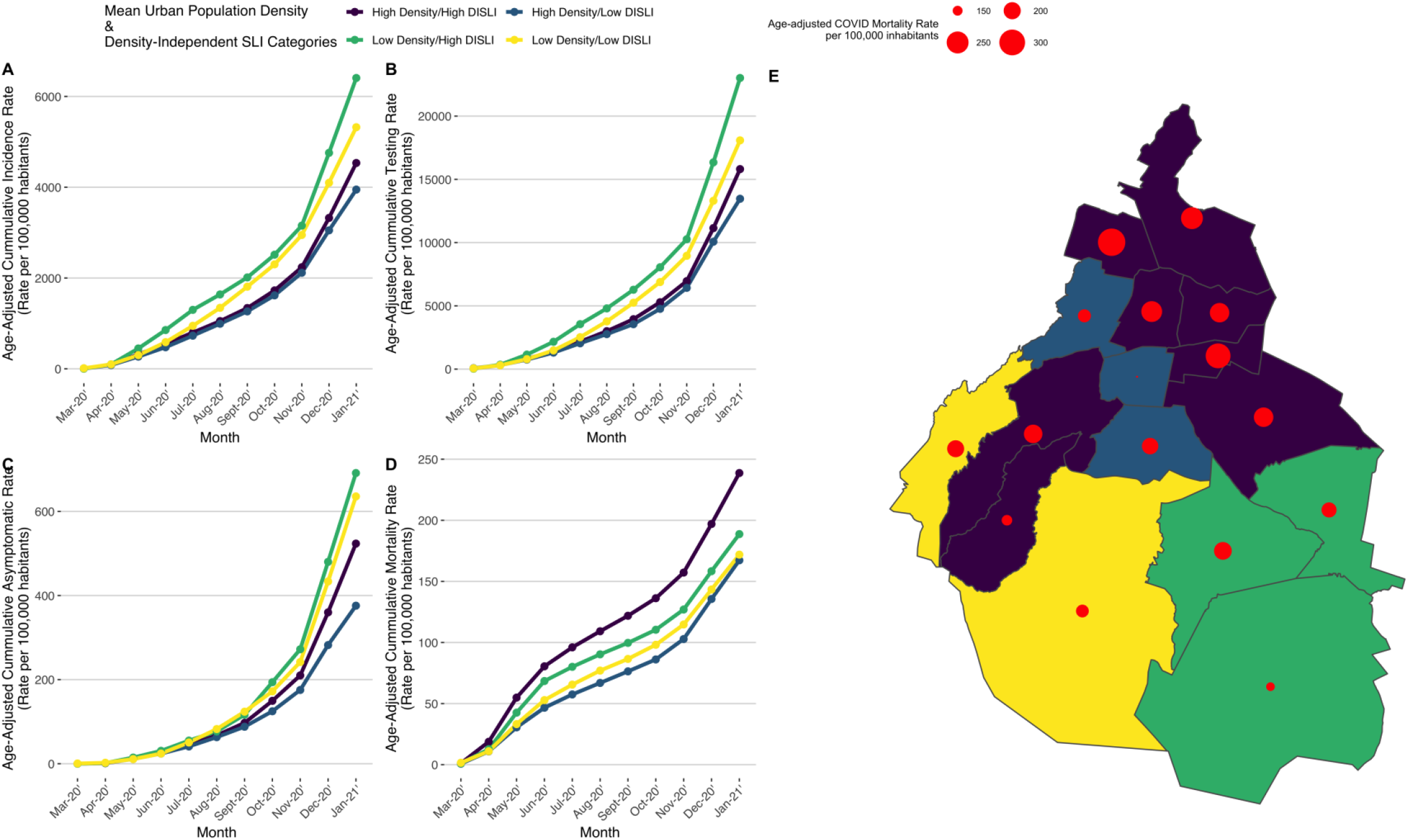
Populational age-adjusted analyses of incidence (A), testing (B), asymptomatic (C) and mortality rates per 100,000 inhabitants (D) according with their mean urban population density and density-independent SLI categories in Mexico City and SLI-terciles categories. Age-adjusted COVID-19 mortality rate according to each individual municipality in Mexico City (D).

### Independent role of individual- and municipal-level factors on COVID-19 outcomes

We sought to investigate whether simultaneous consideration of both individual- and municipal-level factors significantly influence the risk of COVID-19 outcomes. To this end, we fitted multilevel mixed effects Cox and logistic regression models. We observed that unemployed, non-specified workers, home-related workers and retired adults had increased risk for hospitalization, IMV, severe outcomes and mortality compared with HCWs after covariate adjustment. Economically active workers only displayed a risk for IMV, severe outcome and lethality. As expected, the students group had a decreased risk for all evaluated outcomes. Cases treated in a public health-care setting, had a decreased likelihood for hospitalization and IMV requirement, but increased risk of lethality compared with cases treated in private health-care institutions. Furthermore, we observed that subjects living in municipalities with high MUPD had increased risk for all evaluated outcomes. Nevertheless, municipalities classified as High-MUPD/High DISLI displayed the highest risk for hospitalization, IMV, severe outcome and lethality compared with Low MUPD/Low DISLI municipalities (**Figure 2**).

**Figure 2:**
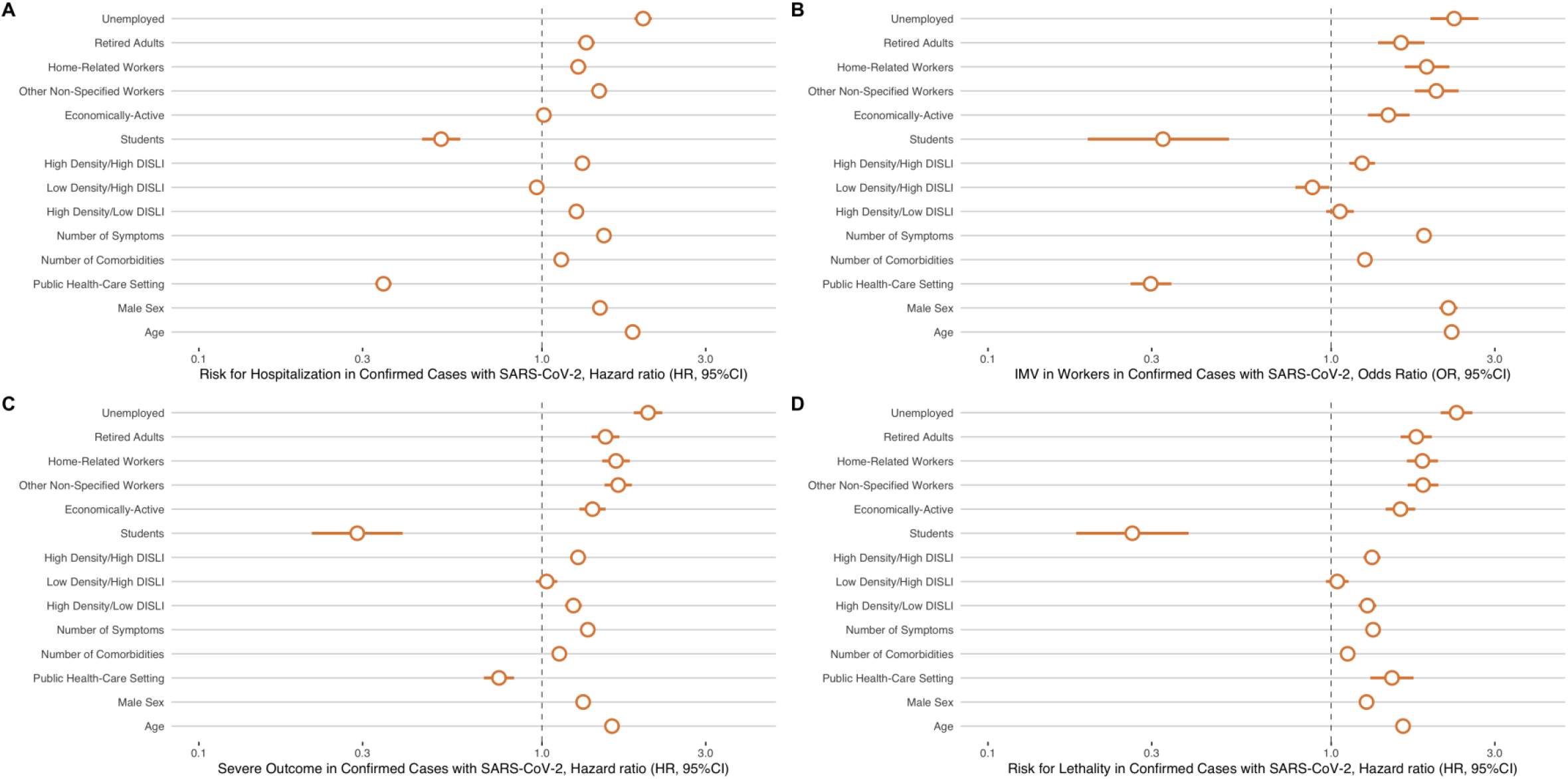
Individual and structural factors associated with hospitalization (A), invasive mechanical ventilation (C), severe outcome (B), and death (D) in patients with a positive RT-PCR for SARS-CoV-2 in Mexico City. Abbreviations: IMV = Invasive Mechanical Ventilation, RT-PCR = Reverse Transcription-Polymerase Chain Reaction, SARS-CoV-2 = Severe Acute Respiratory Coronavirus 2, DISLI = Density Independent Social Lag Index.

**Figure 3.**
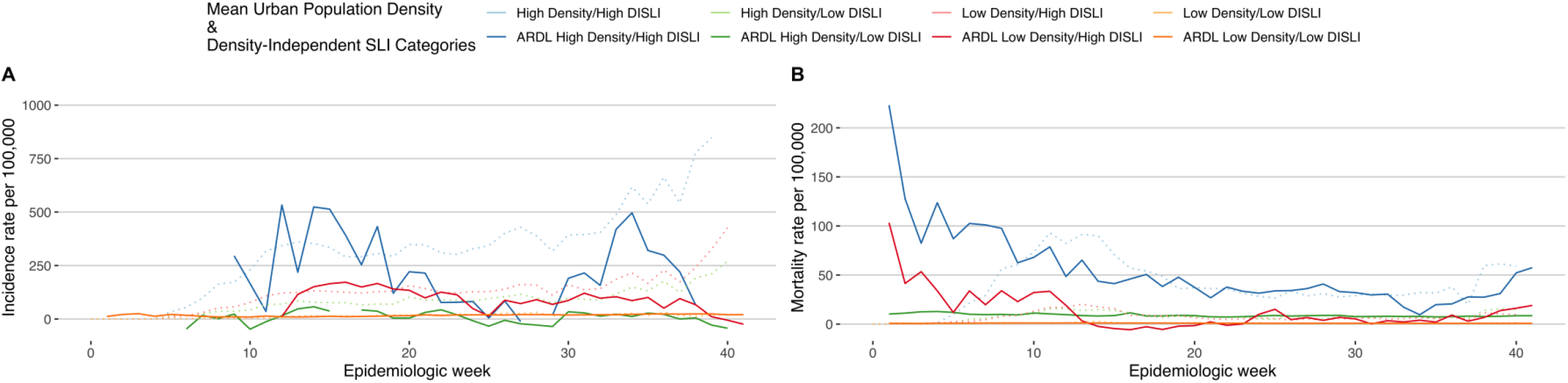
Autoregressive distributed lag (ARDL) models assessing the effect of vehicular mobility restriction policies and its influence on COVID-19 incidence and mortality across mean urban population density and density-independent SLI categories in Mexico City. Abbreviations: DISLI = Density Independent Social Lag Index. Abbreviations:

### Influence of ambulatory setting on adverse outcomes

The burden of COVID-19 in Mexico City caused a more restricted hospital admission criteria, focusing on patients with severe COVID-19. This led to a significant proportion of COVID-19 cases being managed in an outpatient setting. Therefore, we evaluated whether the outpatient management played as an interactor within our working categories on lethality, aggravating the previously identified occupational risk. We observed that unemployed subjects (HR 2.49, 95% CI: 1.87-3.31), retired adults (HR 3.35, 95% CI: 2.56-4.38) and other non-specified workers (HR 1.37, 95% CI: 1.05-1.79) with SARS-CoV2 infection who were treated in an ambulatory setting had increased risk for lethality whilst students (HR 0.04, 95% CI: 0.01-0.14) and economically active workers (HR 0.66, 95% CI: 0.51-0.86) had decreased likelihood for this outcome, adjusted for covariates (**Supplementary Material**). Subsequently, we observed a triple interaction effect for lethality for domestic workers <65 years treated in an ambulatory setting (HR 1.46, 95% CI 1.15-1.87).

### Vehicular mobility has a time-dependent and heterogeneous effect on COVID-19 rates

Vehicular mobility reduction had a heterogeneous behavior across municipalities in Mexico City. At the beginning of the pandemic there were relative decreases in vehicular mobility compared to 2019, but as the pandemic kept going, vehicular mobility increased (**Supplementary Material**). For incidence rates, we observed interaction effects for all MUPD/DISLI categories and vehicular mobility trends. For High MUPD/Low DISLI, we did not observe significant interaction for the first two adjusted splines (decrease <-43.87%) in the model (p=0.33 and p=0.10). For mortality rates, there were also significant interactions for al DISLI/Density quadrants, although the behavior was different than the one observed in incidence rates; significant interactions varied amongst categories (**Supplementary Material**). Severe case rates had significant interactions for High MUPD/Low DISLI (last two splines) with a protective effect for a decrease >27.29% (IRR 0.72, 95%CI 0.53-0.99) and a greater risk when mobility was decreased less than 16.39% (IRR 3.01, 95%CI 1.29-7.02), and for Low MUPD/High DISLI (first spline), observing a protective effect when vehicular mobility was decreased >71.21% (IRR 0.99, 95%CI 0.98-1.00). Hospitalization rates showed significant interactions for all categories as well. All the estimates observed from different rates agreed that, when the interaction was significant, a smaller relative vehicular mobility decrease was associated with an increased risk, except for incidence rate in Low MUPD/Low DISLI quadrant (IRR 0.18, 95%CI, 0.09-0.38). Other predictors, like age, public policy measures, and number of comorbidities were also significant for all models (**Supplementary Material**). We further used the Granger causality tests to assess the temporal relationships between vehicular mobility trends and positivity and mortality rates stratified by either MUPD/DISLI categories or work-group categorization for cointegration. Both mobility and mortality rates were significant predictors of each other both when grouping by worker-group and MUPD/DISLI categories, suggesting a bidirectional relationship. We observed that for the incidence rate, the Granger test was only significant when stated that the incidence rate acted as predictor for changes in vehicular mobility and not the other way around (p=0.006 vs p=0.791) when categorizing by worker-group, but Granger tests when grouping by MUPD/DISLI quadrants had the opposite behavior (p=0.674 vs p=<0.005).

For the ARDL models for individual MUPD/DISLI quadrants with mobility as predictor for incidence, only Low MUPD/Low DISLI had significant short and long term cointegration, and for mortality only Low Density/High DISLI quadrant showed cointegration, even when the rest of the models were significant. When work-group categorization was used as classification criteria, only Students, Economically Active, Homecare Related, and Other Workers showed long-term cointegration, with the rest of the models showing only short term cointegration for the positivity rate. For mortality rates, there was no cointegration for neither Homecare Related and Other Workers, with both long and short term cointegration for the rest of the worker groups (**Supplementary Material**).

### Discrepancies in suspected COVID-19 deaths in Mexico City

As previously mentioned, Mexico City has suffered a mayor hospital reconversion to prioritize attention of patients with COVID-19. We hypothesized that a) this systematic hospital reconversion significantly impacted access to care for several life-threatening non-COVID-19 diseases; b) that this, coupled with the wide-ranging economic disruption caused by the pandemic, produced excess mortality; and c) that this effect was unequally distributed within Mexico City with significant influence by sociodemographic inequalities. Hence, we evaluated excess mortality data attributed to death related and unrelated to COVID-19. We first compared the number of deaths registered in the NESS with death certificates captured by the GCR of Mexico City to assess the magnitude of underreported COVID-19 deaths. The GCR recorded 37,818 deaths with suspected or confirmed COVID-19, of which 20,038 (52.9%) were registered in hospital, 2,454 (6.5%) in an ambulatory setting and 15,326 (40.5%) were unspecified; conversely, the NESS reported 32,076 deaths with confirmed or suspected COVID-19 (29,134 [90.8%] in hospital and 2,942 [9.2%] in an ambulatory setting). Overall, this represents a 15.2% discrepancy compared with suspected COVID-19 deaths registered in the GCR, indicating potential underreported COVID-19 deaths.

### Underreported COVID-19 deaths are dependent on social lag

When adjusting for population, we found that the highest COVID-19 related mortality rates in death certificates occurred in Low MUPD/High DISLI, followed by High MUPD/High DISLI, Low MUPD/Low DISLI and High MUPD/Low DISLI municipalities (**Supplementary Material**). When comparing COVID-19 mortality rates between NESS and the GCR, we found that the discrepancies in recorded deaths were higher in High MUPD/High DISLI municipalities. These differences were steeper in hospital deaths compared with an ambulatory setting. Using negative-binomial models to assess factors related to discrepancies in suspected COVID-19 deaths between the NESS and the GCR we observed that unregistered suspected COVID-19 deaths were associated with increased SLI (IRR 4.46, 95% CI 2.50-7.97). Nevertheless, this association was lost using our DISLI composite variable. Moreover, after grouped for our MUPD/DISLI composite categories, we observed that no municipality displayed an increased rate of unregistered suspected COVID-19 deaths adjusted by the masculinity and dependency indexes.

### Unequal distribution of excess mortality during the COVID-19 pandemic in Mexico City

Regarding age adjusted mortality rates of non-COVID-19 deaths, these have increased during the studied period (March 2020-January-2021, deaths: 82,786; Age-adjusted mortality rate: 9,777 per 100,000 inhabitants) compared to the average mortality rate from 2017 to 2019 (average deaths from 2017 to 2019: 44,707; Age-adjusted mortality rate: 5,259 per 100,000 inhabitants). We observed that 60.5% (n= 41,173) of 2020 Non-COVID-19 deaths were recorded in an ambulatory setting, compared with 34.4% (n=23,428) registered in a hospital and 13.4% (n=3,467) were unspecified. Starting from April-2020, the proportion of non-COVID-19 deaths registered in an ambulatory setting increased compared with a hospital setting until reaching a 2:1 ratio (IQR: 1.29-1.92); we observed no significant differences in this ratio across MUPD/DISLI categories. Conversely, the rate of ambulatory to hospital deaths by COVID-19 showed that these occurred in substantial proportion within hospitals (**Supplementary Material**).

Finally, we observed an excess mortality (Overall age-adjusted excess mortality: 4,518 per 100,000 inhabitants) of non-COVID-19 deaths which had the highest peak in May-2020 and was consistently higher in Moderate and High-DISLI municipalities. Overall, we observed a non-linear association for the DISLI to predict higher excess mortality of non-COVID-19 deaths (**Supplementary Material**). Using negative-binomial regression, we identified that compared to Low-MUPD/Low DISLI municipalities, High-MUPD/High DISLI (IRR 3.88, 95%CI 3.36-4.47), Low-MUPD/High DISLI (IRR 1.33, 95%CI 1.15-1.54) and High MUPD/Low DISLI (IRR 1.24, 95%CI 1.07-1.43) had increased excess non-COVID-19 deaths adjusted by the masculinity and dependency indexes and the ambulatory to hospitalities deaths ratio (**Figure 4**).

**Figure 4:**
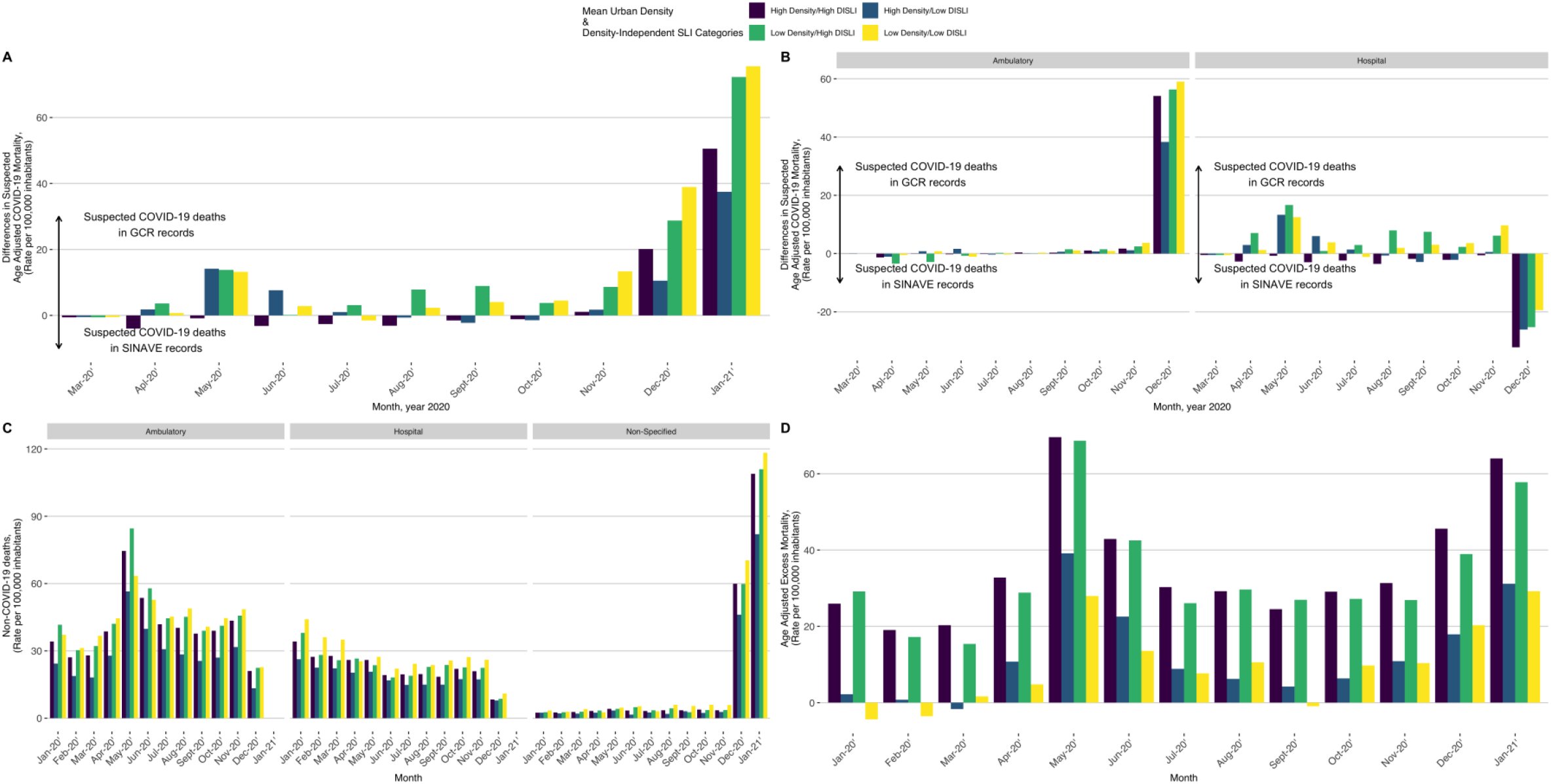
Differences in mortality rate of suspected COVID-19 deaths between the NESS and GCR (A) stratified by ambulatory or hospital setting (B). Mortality rate of non-COVID-19 deaths by place of death (C) and excess mortality compared with year 2019 (D) across mean urban population density and density-independent SLI categories in Mexico City.

## DISCUSSION

In this work, we evaluated whether sociodemographic and structural factors at an individual and municipal level influenced COVID-19 outcomes during the pandemic in Mexico City. We detected vulnerable individuals whose working status likely prevented adherence to social distancing mandates. Furthermore, we observed that the mix of high MUPD and marginalization, as measured by DISLI, leads both to higher mortality rates in the community and higher risk for adverse outcomes for individuals living in such communities. Additionally, social-distancing policies as proxied by reduced vehicular mobility, had differential impacts across municipalities depending on social lag, suggesting that such policies have helped to decrease the impact of the pandemic, but had a lesser effect on halting community-level transmission in socially vulnerable municipalities. Finally, we report discrepancies between death certificates for suspected COVID-19 deaths and the NESS, which are in line with reports from other countries (4,6,7,12–14). Nevertheless, the dependency on DISLI and MUPD for both COVID- and non-COVID-related excess deaths shows the extent to which the COVID-19 pandemic has affected the most vulnerable sectors of society. These findings indicate that healthcare policies focused on attending severe COVID-19 cases such as hospital reconversion strategies had negative externalities, particularly on marginalized municipalities in Mexico City.

Our results support previous evidence which show that densely populated communities living in marginalization have increased risk of sustained community transmission of various infectious diseases, including SARS-CoV-2 (39–41). Furthermore, densely populated, and marginalized municipalities in Mexico City had sustained community-level transmission and a considerable number of asymptomatic cases despite mitigation measures, which notably had different effects modified by municipal social lag. Overall, our results highlight that inequalities at the individual and municipal level had a significant impact on the course of the COVID-19 pandemic in Mexico City. The precise *pathways of embodiment* and the *processes of social determination* through which mean urban density and marginalization lead to higher rates of COVID-19 related adverse outcomes requires further quantitative and qualitative research. Broadly, they can be conceptualized as structural factors and factors related to healthcare provision. The map of COVID-19 mortality and excess mortality is remarkably similar to the map of lower life expectancy, perhaps suggesting that structural factors which increased risk for transmission and adverse outcomes play a larger role than those related to healthcare provision. This is relevant, as life expectancy is generally thought to be more closely related to standards of living than to healthcare provision (19).

Local government public policy has been directed at tackling inequities in dealing with the COVID-19 pandemic, with actions including prioritizing case-finding in marginalized neighborhoods, providing stipends for confirmed cases to facilitate self-isolation, as well as hospital reconversion to increase capacity for healthcare delivery during pandemic stress periods (42). Although these actions have probably attenuated the impact of COVID-19 on vulnerable communities, our results show that the pandemic has had differential impacts along socioeconomic lines. This is clear when considering that social isolation policies had a heterogeneous impact on the way the pandemic has hit different municipalities across Mexico City. What we observed from the behavior of vehicular mobility is that there is indeed a key cutoff where vehicular mobility decreases influences COVID-19 rates in Mexico City. As shown by our results, untargeted strategies to reduce urban mobility may not be effective for all areas of Mexico City, considering the influence of marginalization and MUPD may inform future public health policy measures. To be effective in halting the impact of the COVID-19 pandemic, these measures need to be adapted to the specific urban characteristics of these communities.

Mexico City is one of the most densely populated metropolises in Latin America, but also one of the cities with the highest inequality in income distribution (43). These geographic and economic disparities are translated into social and structural deficiencies which have impacted healthcare access and quality of care even before the onset of the COVID-19 pandemic. It has been proposed that social lag is a significant limiting factor for access to sufficient healthcare services (44). In our results, other vulnerable groups such as retired adults, domestic workers and unemployed individuals showed a greater predisposition to COVID-19 mortality. These subjects could be part of the informal economy and live in conditions which prevent them guaranteed access to healthcare services; despite efforts by the local authorities to provide financial support to promote self-isolation, these individuals still likely required to work for economical sustenance after the relief of social distancing mandates (45). Evidence supports that living with economically active workers increases the risk of contracting SARS-CoV2 infection, particularly in older adults, which may partly explain the risk of transmission within these groups (46). Systematic policies are required to guarantee equitable access to health services, prompt detection of SARS-CoV2 infection, and sufficient stipends which facilitate self-isolation to prevent SARS-CoV-2 infection for vulnerable groups.

Finally, our findings also reveal that excess mortality from non-COVID-19 deaths increased considerably, most notably in outpatient settings. Excess mortality was predicted as an unwanted effect of the pandemic, which varies according to each country (47–50). Excess mortality in Mexico City may result from deficiencies within the infrastructure of healthcare systems along with a fragmented hospital framework which complicates interoperability (50,51). These factors coexisted within Mexico City and overall in Mexico well before the COVID-19 pandemic (20,52); furthermore, insufficient number of HCWs and the burden that this sector had during the pandemic created a triple overload for the Mexican healthcare system (31,53,54). As a result of this urgent need, local authorities sought to create a strategy based on hospitals’ reconversion and modified patient admission criteria to prioritize patients with severe COVID-19. This systematic hospital reconversion led to undesired negative externalities, where a vulnerable sector of the population living in marginalized municipalities had to restrict attention for non-COVID-19 related medical ailments and were less likely to seek urgent medical attention in an emergency setting. Furthermore, it is likely that the effects of the economic upheaval caused by the pandemic has had a larger impact precisely on marginalized populations, creating further difficulty in meeting essential needs and thus leading to higher excess mortality. This excess mortality could also be accounted for by unregistered COVID-19 deaths, indirect effects of the pandemic or increased mobilization of medical urgencies coming from neighboring states seeking attention given potential unavailability of medical services. Further work is required to elucidate underlying causes of excess deaths to truly assess the negative externalities of hospital reconversion during the COVID-19 pandemic in Mexico City.

Our study brings novel insights of the COVID-19 pandemic in Mexico City. The integration of several population-based datasets provides information related to the role of individual and municipality-wide structural and social determinants that interplayed and ultimately modified the course of the pandemic. Nevertheless, some limitations should be acknowledged. All the estimations made in our population analyses were based on projections of the economic population of the first quarter of 2020 and may not capture the dynamics around employment occupation and changes in population structure within the study period. Furthermore, since there are no data on the demographic structure of the working categories, we were unable to perform age-adjustment for this section of the analysis. Second, our categorization by MUPD/DISLI categories was based solely on Mexico City, which may not represent the country’s socio-economic disparities or heterogeneity within smaller communities or individual socioeconomic levels. Third, the NESS may not fully capture patients with mild and asymptomatic SARS-CoV-2 infection, which represents a bias towards cases with moderate and high risk for COVID-19 complications. Finally, excess mortality data does not report disaggregated etiology of non-COVID-19 deaths, which prevents further assessment of areas which may have required further attention during the pandemic to prevent excess deaths.

In summary, individual- and municipal-level socioeconomic and structural factors played a significant role in shaping the course of the COVID-19 pandemic in Mexico City. Notably, despite higher incidence of SARS-CoV-2 infection in economically active workers, a higher burden of adverse COVID-19 outcomes was observed in non-specified workers, retired adults, home related, and unemployed workers. MUPD independent municipal social lag was a risk factor for COVID-19 adverse outcomes and its effect was modified by urban population density; furthermore, DISLI also influenced the protective effect of vehicular mobility reduction on Mexico City. Finally, excess mortality was dependent on social lag, whilst underreport of COVID-19 deaths is related to MUPD. The inequalities highlighted by the COVID-19 pandemic call for urgent actions to reduce socioeconomic disparities, increase healthcare access and promote healthier lifestyles in Mexico City and beyond.

## Data Availability

All data sources and R code are available for reproducibility of results at: https://github.com/oyaxbell/covid_inequality_cdmx/

## ACKNOWLEDGMENTS

NEAV, JPBL, AVV, AMS, and CAFM are enrolled at the PECEM program of the Faculty of Medicine at UNAM. NEAV, JPBL and AVV are supported by CONACyT. The authors would like to acknowledge the invaluable work of all of Mexico’s healthcare community in managing the COVID-19 pandemic. Their participation in the COVID-19 surveillance program alongside with open data sharing has made this work a reality, we are thankful for your effort. This work is registered at Instituto Nacional de Geriatría under project number DI-PI-005/2021.

## AUTHORS’ CONTRIBUTIONS

Research idea and study design NEAV, LFC, JPA, OYBC; data acquisition: NEAV, LFC, JPA, AGD, OYBC; data analysis/interpretation: NEAV, LFC, JPA, JMG, AKG, AVV, AGD, CAFM, AMS, ECG, JPBL, MVR, JSM, MEJM, LMGR, OYBC; statistical analysis: NEAV, LFC, JPA, OYBC; manuscript drafting: NEAV, LFC, JPA, JMG, LMGR, OYBC; supervision or mentorship: LMGR, OYBC. Each author contributed important intellectual content during manuscript drafting or revision and accepts accountability for the overall work by ensuring that questions about the accuracy or integrity of any portion of the work are appropriately investigated and resolved.

