## Supplementary Material for "Comprehensive evaluation of the impact of sociodemographic inequalities on adverse outcomes and excess mortality during the COVID-19 pandemic in Mexico City"

### SUPPLEMENTARY METHODS

#### COVID-19 case definition

Criteria for suspected COVID-19 case in Mexico included having at least two of three signs/symptoms (cough, fever, or headache) plus at least one other (dyspnea, arthralgias, myalgias, sore throat, rhinorrhea, conjunctivitis, or chest pain) in the last 7 days. This operational definition was changed on August 24, 2020 to increase sensitivity: at least one of four signs/symptoms (cough, fever, dyspnea, or headache), plus at least one other (myalgias, arthralgias, sore throat, chills, chest pain, rhinorrhea, anosmia, dysgeusia, or conjunctivitis) in the last 10 days.

#### National Epidemiological Surveillance Study dataset

All demographic and health data were collected and uploaded to the NESS database by health-care personnel from each corresponding health-care facility. Available variables include age, sex, nationality, state and municipality where the case was detected, immigration status as well as identification of individuals who self-identify as indigenous. Health information includes the status of diabetes, obesity, chronic obstructive pulmonary disease (COPD), immunosuppression, pregnancy, arterial hypertension, cardiovascular disease, chronic kidney disease (CKD), and asthma. Evaluated symptoms included fever, cough, odynophagia, dyspnea, irritability, diarrhea, chest pain, shivering, headache, myalgia, arthralgia, malaise, rhinorrhea, polypnea, vomiting, abdominal pain, conjunctivitis, cyanosis, and sudden onset of symptoms.

#### Working group classification according to the National Survey of Employment and Occupation

The National Survey of Employment and Occupation (ENEO) considers as economically-active employers to those whom self-reported as merchants, transport workers, laborers, farmers, teachers and managers/owners of a business at the moment of evaluation. The health-care workers were classified according to those evaluated subjects whom self-reported as physicians, nurses, dentists, laboratorians and other-related HCWs. The group of unemployed subjects included those who reported as not being hired or affiliated in any working area or who did not specify any previously mentioned working profession at the moment of evaluation. Finally, we assign a working category group for the students, retired workers, home-care related workers and other non-specified workers.

### Social lag index components

The SLI is composite of several components that are measured to estimate social disadvantage and structural inequality at a municipal level based on population census data from the Mexican National Evaluation Council (CONEVAL); SLI is a principal component score which comprises percentages of literacy, access to basic education, healthcare services, living conditions including drainage, dirt floor, access to water, electricity and electrical appliances for each Mexican municipality. Although CONEVAL proposes a categorization by quintiles to assess social lag by municipality for the entire country, Mexico City represents an exception. The municipalities of Mexico City are categorized as a low lag compared to the rest of the country. To analyze the specific case of the Mexico City, we categorized by terciles of SLI according to their distribution municipality assigned to the municipality of residence for each evaluated case in the NESS.

### Death certificates of Mexico City General Civil Registry

It contains the information on the death certificates registered before the Civil Registry of Mexico City in 2017, 2018, 2019, and 2020. The variables included in the GCR dataset are the date of death, the municipality where the death was registered, the place of the death (outpatient, hospital, or unknown), and if it was due to suspected COVID-19 or non-COVID-19 cause.

### SUPPLEMENTARY TABLES

**Supplementary Table 1:** Estimated population during the first trimester of 2020 of economically capable population >15 years estimated by the National Survey of Employment and Occupation. *: HCWS= This group included subjects whose occupations were reported as physicians, nurses, dentists, laboratory personnel and other involved HCWs (Secretaria de Salud. Sistema de Información de Salud [Internet]. 2019. Available from: http://sinaiscap.salud.gob.mx:8080/DGIS/). Economically-Active workers were defined as those whom self-reported as managers/owners of a business, teachers, laborers, employers, merchants, transport workers and farmers

| **Working category** | **Estimated population** |
|---|---|
| Economically-Active workers | 4,045,727 |
| Retired workers | 468,737 |
| Home-care related workers | 1,287,289 |
| Students | 767,782 |
| Other unspecified non-active subjects | 316,058 |
| Health-care workers | 161,580 |
| Unemployed workers | 208,130 |
| Overall working capable population | 7,255,303 |

**Supplementary table 3:** Characteristics of suspected COVID-19 cases as 31th of January of 2021 divided by working group categories in Mexico City. *Abbreviations*: COPD: Chronic obstructive pulmonary disease; HIV/AIDS: Human immunodeficiency virus and/or acquired immunodeficiency syndrome; CKD: Chronic kidney disease, CVD: cardiovascular disease.

| | All population | | HCWs | | Students | | Economically-Active | | Other Non-Specified workers | | Home-Related workers | | Unemployed | | Retired Adults | | P values |
|---|---|---|---|---|---|---|---|---|---|---|---|---|---|---|---|---|---|
| Age [years] | 40.70 | (±16.79) | 38.83 | (±11.43) | 18.02 | (±8.07) | 41.05 | (±13.00) | 40.75 | (±16.87) | 50.55 | (±17.11) | 43.42 | (±18.67) | 68.02 | (±10.62) | <0.001 |
| Female sex [%] | 763753 | (76.71%) | 61086 | (64.82%) | 81290 | (52.43%) | 319229 | (44.37%) | 73198 | (41.97%) | 199949 | (94.21%) | 16476 | (31.34%) | 12525 | (31.20%) | <0.001 |
| Positivity [%] | 419666 | (42.15%) | 27520 | (29.20%) | 40882 | (26.37%) | 199060 | (27.67%) | 52399 | (30.04%) | 69931 | (32.95%) | 14995 | (28.52%) | 14879 | (37.07%) | <0.001 |
| Asymptomatics [%] | 46729 | (4.69%) | 1241 | (1.32%) | 6954 | (4.49%) | 22369 | (3.11%) | 5882 | (3.37%) | 7876 | (3.71%) | 1096 | (2.08%) | 1311 | (3.27%) | <0.001 |
| Mortality [%] | 31820 | (3.20%) | 607 | (0.64%) | 80 | (0.05%) | 9547 | (1.33%) | 5143 | (2.95%) | 6986 | (3.29%) | 3641 | (6.93%) | 5816 | (14.49%) | <0.001 |
| Hospitalization [%] | 82342 | (8.27%) | 3408 | (3.62%) | 1709 | (1.10%) | 27069 | (3.76%) | 14692 | (8.42%) | 16404 | (7.73%) | 8376 | (15.93%) | 10684 | (26.62%) | <0.001 |
| Clinical pneumonia [%] | 67105 | (6.74%) | 4587 | (4.87%) | 1295 | (0.84%) | 24207 | (3.36%) | 10388 | (5.96%) | 13179 | (6.21%) | 5591 | (10.64%) | 7858 | (19.58%) | <0.001 |
| Severe outcome [%] | 21253 | (2.13%) | 540 | (0.57%) | 60 | (0.04%) | 6632 | (0.92%) | 3344 | (1.92%) | 4636 | (2.18%) | 2302 | (4.38%) | 3739 | (9.32%) | <0.001 |
| Mechanical ventilation [%] | 12651 | (1.27%) | 381 | (0.40%) | 98 | (0.06%) | 4473 | (0.62%) | 2266 | (1.30%) | 2563 | (1.21%) | 1122 | (2.13%) | 1748 | (4.35%) | <0.001 |
| <7 days since beginning of symptoms [%] | 224609 | (22.56%) | 13292 | (14.10%) | 20459 | (13.20%) | 109551 | (15.23%) | 26582 | (15.24%) | 37733 | (17.78%) | 8256 | (15.70%) | 8736 | (21.76%) | <0.001 |
| Subjects with diabetes [%] | 120198 | (12.07%) | 5278 | (5.60%) | 799 | (0.52%) | 49856 | (6.93%) | 13536 | (7.76%) | 33854 | (15.95%) | 5883 | (11.19%) | 10992 | (27.38%) | <0.001 |
| Hypertension [%] | 161840 | (16.26%) | 8677 | (9.21%) | 922 | (0.59%) | 65081 | (9.05%) | 17763 | (10.18%) | 46813 | (22.06%) | 7203 | (13.70%) | 15381 | (38.32%) | <0.001 |
| Asthma [%] | 31476 | (3.16%) | 3772 | (4.00%) | 4728 | (3.05%) | 13941 | (1.94%) | 3258 | (1.87%) | 4387 | (2.07%) | 812 | (1.54%) | 578 | (1.44%) | <0.001 |
| COPD [%] | 9255 | (0.93%) | 366 | (0.39%) | 152 | (0.10%) | 2703 | (0.38%) | 1103 | (0.63%) | 2851 | (1.34%) | 667 | (1.27%) | 1413 | (3.52%) | <0.001 |
| HIV/AIDS [%] | 5055 | (0.51%) | 278 | (0.29%) | 240 | (0.15%) | 2697 | (0.37%) | 840 | (0.48%) | 487 | (0.23%) | 331 | (0.63%) | 182 | (0.45%) | <0.001 |
| CVD [%] | 16469 | (1.65%) | 1072 | (1.14%) | 623 | (0.40%) | 5889 | (0.82%) | 2005 | (1.15%) | 3889 | (1.83%) | 918 | (1.75%) | 2073 | (5.16%) | <0.001 |
| CKD [%] | 10836 | (1.09%) | 382 | (0.41%) | 322 | (0.21%) | 3633 | (0.50%) | 1420 | (0.81%) | 2712 | (1.28%) | 1038 | (1.97%) | 1329 | (3.31%) | <0.001 |
| Obesity [%] | 141479 | (14.21%) | 12419 | (13.18%) | 6411 | (4.13%) | 71952 | (10.00%) | 15837 | (9.08%) | 26128 | (12.31%) | 4333 | (8.24%) | 4399 | (10.96%) | <0.001 |
| Smoking stat1us [%] | 159147 | (15.99%) | 8975 | (9.52%) | 8013 | (5.17%) | 95016 | (13.21%) | 19205 | (11.01%) | 15127 | (7.13%) | 8780 | (16.70%) | 4031 | (10.04%) | <0.001 |
| Pregnant women [%] | 7465 | (0.75%) | 298 | (0.32%) | 350 | (0.23%) | 1784 | (0.25%) | 774 | (0.44%) | 4174 | (1.97%) | 82 | (0.16%) | 3 | (0.01%) | <0.001 |
| Indigenous [%] | 5487 | (0.55%) | 297 | (0.32%) | 356 | (0.23%) | 2772 | (0.39%) | 711 | (0.41%) | 977 | (0.46%) | 227 | (0.43%) | 147 | (0.37%) | <0.001 |

**Supplementary Table 4:** Characteristics of suspected COVID-19 cases as 31th of January of 2021 by MUPD/DISLI quadrants in Mexico City. *Abbreviations*: COPD: Chronic obstructive pulmonary disease; HIV/AIDS: Human immunodeficiency virus and/or acquired immunodeficiency syndrome; CKD: Chronic kidney disease, CVD: cardiovascular disease.

| | General | | High MUPD/High DISLI | | High MUPD /Low DISLI | | Low MUPD / High DISLI | | Low MUPD /Low DISLI | | P-value |
|---|---|---|---|---|---|---|---|---|---|---|---|
| Age [years] | 40.70 | ±16.79 | 41.13 | ±16.79 | 41.77 | ±16.81 | 39.06 | ±16.81 | 39.14 | ±16.62 | <0.001 |
| Female sex [%] | 763753 | (52.74%) | 470002 | (52.64%) | 103385 | (52.77%) | 174173 | (52.92%) | 16193 | (53.66%) | <0.001 |
| Positivity [%] | 419666 | (28.98%) | 258929 | (29.00%) | 58678 | (29.95%) | 92779 | (28.19%) | 9280 | (30.75%) | <0.001 |
| Asymptomatics [%] | 46729 | (3.23%) | 29747 | (3.33%) | 5420 | (2.77%) | 10272 | (3.12%) | 1290 | (4.27%) | <0.001 |
| Mortality [%] | 31820 | (2.20%) | 22550 | (2.53%) | 4716 | (2.41%) | 4101 | (1.25%) | 453 | (1.50%) | <0.001 |
| Hospitalization [%] | 82342 | (5.69%) | 55759 | (6.24%) | 13362 | (6.82%) | 11910 | (3.62%) | 1311 | (4.34%) | <0.001 |
| Clinical pneumonia [%] | 67105 | (4.63%) | 45254 | (5.07%) | 10133 | (5.17%) | 10728 | (3.26%) | 990 | (3.28%) | <0.001 |
| Severe outcome [%] | 21253 | (1.47%) | 14938 | (1.67%) | 3207 | (1.64%) | 2780 | (0.84%) | 328 | (1.09%) | <0.001 |
| IMV[%] | 12651 | (0.87%) | 8480 | (0.95%) | 1918 | (0.98%) | 2066 | (0.63%) | 187 | (0.62%) | <0.001 |
| <7 days since beginning of symptoms [%] | 224609 | (15.51%) | 131788 | (14.76%) | 33238 | (16.97%) | 56959 | (17.31%) | 2624 | (8.70%) | <0.001 |
| Subjects with diabetes [%] | 120198 | (8.30%) | 77401 | (8.67%) | 15814 | (8.07%) | 25158 | (7.64%) | 1825 | (6.05%) | <0.001 |
| Hypertension [%] | 161840 | (11.18%) | 104329 | (11.68%) | 23372 | (11.93%) | 31881 | (9.69%) | 2258 | (7.48%) | <0.001 |
| Asthma [%] | 31476 | (2.17%) | 19321 | (2.16%) | 6063 | (3.09%) | 5760 | (1.75%) | 332 | (1.10%) | <0.001 |
| COPD [%] | 9255 | (0.64%) | 6240 | (0.70%) | 1455 | (0.74%) | 1425 | (0.43%) | 135 | (0.45%) | <0.001 |
| HIV/AIDS [%] | 5055 | (0.35%) | 3309 | (0.37%) | 931 | (0.48%) | 765 | (0.23%) | 50 | (0.17%) | <0.001 |
| CVD [%] | 16469 | (1.14%) | 10737 | (1.20%) | 2703 | (1.38%) | 2827 | (0.86%) | 202 | (0.67%) | <0.001 |
| CKD [%] | 10836 | (0.75%) | 7164 | (0.80%) | 1608 | (0.82%) | 1913 | (0.58%) | 151 | (0.50%) | <0.001 |
| Obesity [%] | 141479 | (9.77%) | 90169 | (10.10%) | 20302 | (10.36%) | 28816 | (8.76%) | 2192 | (7.26%) | <0.001 |
| Smoking stat1us [%] | 159147 | (10.99%) | 97795 | (10.95%) | 25645 | (13.09%) | 34217 | (10.40%) | 1490 | (4.94%) | <0.001 |
| Pregnant women [%] | 7465 | (0.52%) | 3983 | (0.45%) | 862 | (0.44%) | 2340 | (0.71%) | 280 | (0.93%) | <0.001 |
| Indigenous [%] | 5487 | (0.38%) | 3163 | (0.35%) | 574 | (0.29%) | 1693 | (0.51%) | 57 | (0.19%) | <0.001 |

**Supplementary Table 5:** Poisson mixed-effect models estimates for incidence, mortality, severe case, and hospitalization rates.

| | Incidence | | | Mortality | | | Severe case | | | Hospitalization | | |
|---|---|---|---|---|---|---|---|---|---|---|---|---|
| | Estimate | 95%IC | *p* | Estimate | 95%IC | *p* | Estimate | 95%IC | *p* | Estimate | 95%IC | *p* |
| Intercept | 0.00 | (0.00-0.00) | <0.005 | 0.00 | (0.00-0.00) | <0.005 | 0.00 | (0.00-0.00) | <0.005 | 0.00 | (0.00-0.00) | <0.005 |
| High Density/Low DISLI | 1.34 | (0.67-2.69) | 0.41 | 0.86 | (0.41-1.81) | 0.70 | 1.13 | (0.53-2.39) | 0.75 | 0.88 | (0.52-1.49) | 0.64 |
| Low Density/High DISLI | 1.30 | (0.66-2.55) | 0.45 | 0.44 | (0.22-0.85) | 0.02 | 0.53 | (0.24-1.17) | 0.11 | 0.58 | (0.35-0.98) | 0.04 |
| Low Density/Low DISLI | 5.23 | (1.61-17.04) | 0.01 | 0.72 | (0.19-2.70) | 0.62 | 2.10 | (0.63-7.01) | 0.23 | 0.99 | (0.40-2.47) | 0.99 |
| M1 (cut-off: -71.21%) | 1.00 | (1.00-1.62) | 0.11 | 1.02 | (1.01-1.02) | <0.005 | 1.01 | (1.01-1.02) | <0.005 | 1.02 | (1.01-1.02) | <0.005 |
| M2 (cut-off: -43.87%) | 0.98 | (0.98-0.99) | <0.005 | 0.95 | (0.93-0.96) | <0.005 | 0.96 | (0.94-0.98) | <0.005 | 0.96 | (0.95-0.97) | <0.005 |
| M3 (cut-off: -27.29%) | 1.11 | (1.08-1.15) | <0.005 | 1.33 | (1.21-1.47) | <0.005 | 1.21 | (1.07-1.36) | <0.005 | 1.17 | (1.10-1.24) | <0.005 |
| M4 (cut-off: -16.39%) | 0.79 | (0.74-0.84) | <0.005 | 0.58 | (0.46-0.72) | <0.005 | 0.66 | (0.51-0.87) | <0.005 | 0.76 | (0.67-0.87) | <0.005 |
| Age | 1.00 | (1.00-33.97) | <0.005 | 1.07 | (1.07-1.07) | <0.005 | 1.06 | (1.06-1.06) | <0.005 | 1.04 | (1.04-1.04) | <0.005 |
| Period | 1.07 | (1.00-1.15) | 0.04 | 0.65 | (0.55-0.77) | <0.005 | 0.73 | (0.58-0.92) | 0.01 | 0.80 | (0.71-0.89) | <0.005 |
| Number of comorbidities | 1.05 | (1.04-1.05) | <0.005 | 1.30 | (1.29-1.32) | <0.005 | 1.32 | (1.31-1.34) | <0.005 | 1.29 | (1.29-1.30) | <0.005 |
| High Density/Low DISLI*M1 | 1.00 | (1.00-1.00) | 0.33 | 1.00 | (0.99-1.00) | 0.55 | 1.00 | (0.99-1.01) | 0.63 | 1.00 | (0.99-1.00) | 0.02 |
| High Density/Low DISLI*M2 | 1.01 | (1.00-1.02) | 0.10 | 1.04 | (1.01-1.08) | 0.02 | 1.04 | (0.99-1.09) | 0.12 | 1.05 | (1.03-1.07) | <0.005 |
| High Density/Low DISLI*M3 | 0.91 | (0.86-0.97) | <0.005 | 0.75 | (0.59-0.96) | 0.02 | 0.72 | (0.53-0.99) | 0.04 | 0.74 | (0.65-0.85) | <0.005 |
| High Density/Low DISLI*M4 | 1.42 | (1.16-1.73) | <0.005 | 2.36 | (1.22-4.58) | 0.01 | 3.01 | (1.29-7.02) | 0.01 | 2.49 | (2.75-3.54) | <0.005 |
| Low Density/High DISLI*M1 | 1.00 | (0.99-1.00) | <0.005 | 0.98 | (0.98-0.99) | <0.005 | 0.99 | (0.98-1.00) | 0.01 | 0.99 | (0.98-0.99) | <0.005 |
| Low Density/High DISLI*M2 | 1.02 | (1.01-1.03) | <0.005 | 1.03 | (0.99-1.07) | 0.13 | 1.02 | (0.97-1.07) | 0.45 | 1.02 | (1.00-1.04) | 0.04 |
| Low Density/High DISLI*M3 | 0.91 | (0.86-0.97) | <0.005 | 0.92 | (0.73-1.16) | 0.47 | 0.98 | (0.73-1.30) | 0.86 | 0.94 | (0.83-1.07) | 0.37 |
| Low Density/High DISLI*M4 | 1.25 | (1.11-1.41) | <0.005 | 1.10 | (0.66-1.86) | 0.71 | 0.97 | (0.52-1.83) | 0.93 | 1.05 | (0.80-1.39) | 0.71 |
| Low Density/Low DISLI*M1 | 1.01 | (1.00-1.01) | <0.005 | 0.98 | (0.97-1.00) | 0.01 | 1.00 | (0.98-1.01) | 0.65 | 0.99 | (0.98-0.99) | <0.005 |
| Low Density/Low DISLI*M2 | 0.94 | (0.92-0.96) | <0.005 | 1.14 | (1.03-1.26) | 0.01 | 1.05 | (0.95-1.16) | 0.31 | 1.07 | (1.02-1.12) | <0.005 |
| Low Density/Low DISLI*M3 | 1.65 | (1.39-1.97) | <0.005 | 0.58 | (0.25-1.35) | 0.20 | 0.94 | (0.38-2.33) | 0.90 | 0.79 | (0.60-1.03) | 0.08 |
| Low Density/Low DISLI*M4 | 0.18 | (0.09-0.38) | <0.005 | 1.98 | (0.06-62.06) | 0.70 | 0.17 | (0.00-14.60) | 0.43 | 1.29 | (0.51-3.30) | 0.59 |

### SUPPLEMENTARY FIGURES

**Supplementary figure 1:** COVID-19 rates according to occupation categories using as denominator 2020 reports of occupation by INEGI.

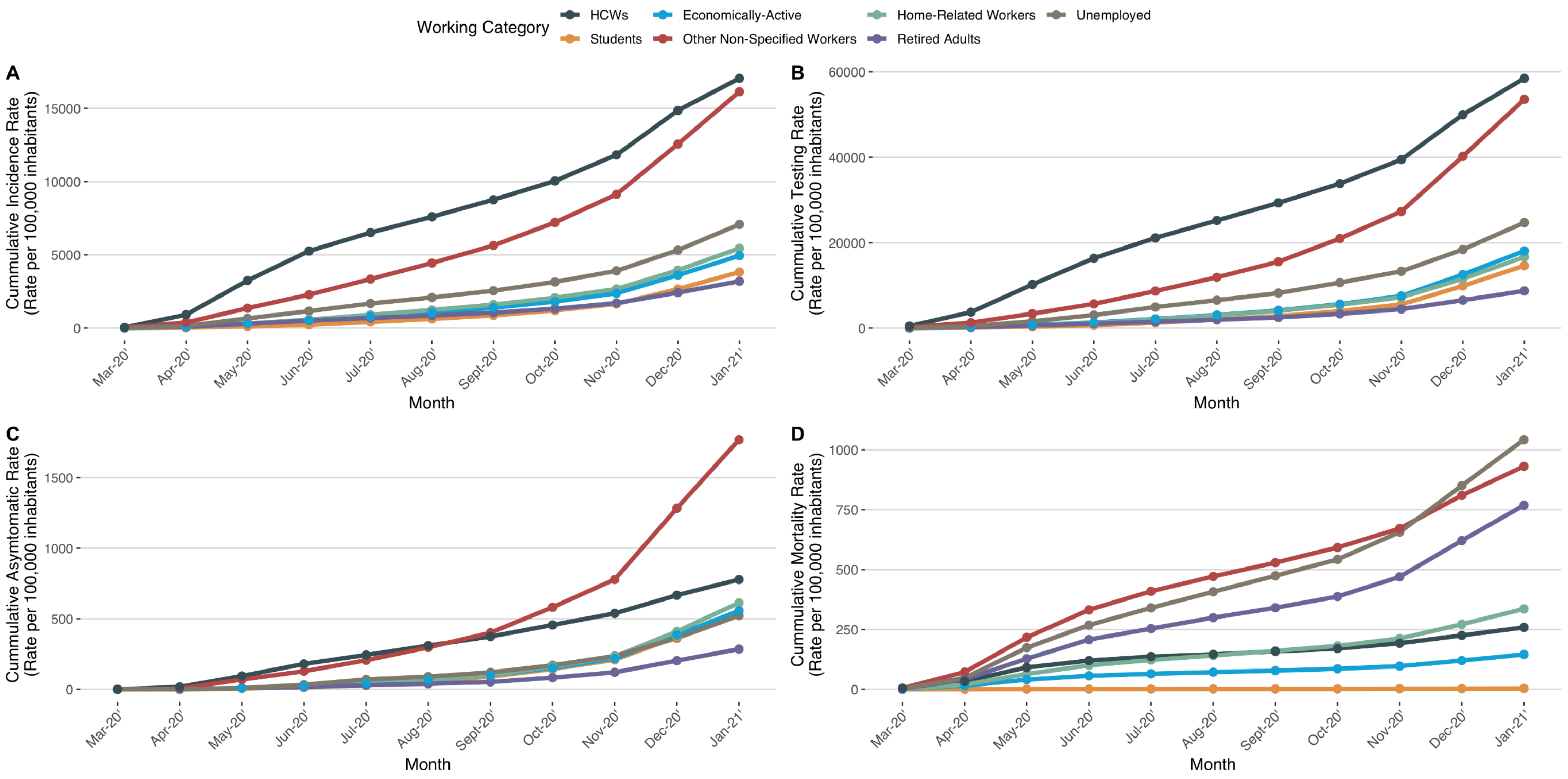

**Supplementary figure 2:** Timeline of COVID-19 pandemic in Mexico City.

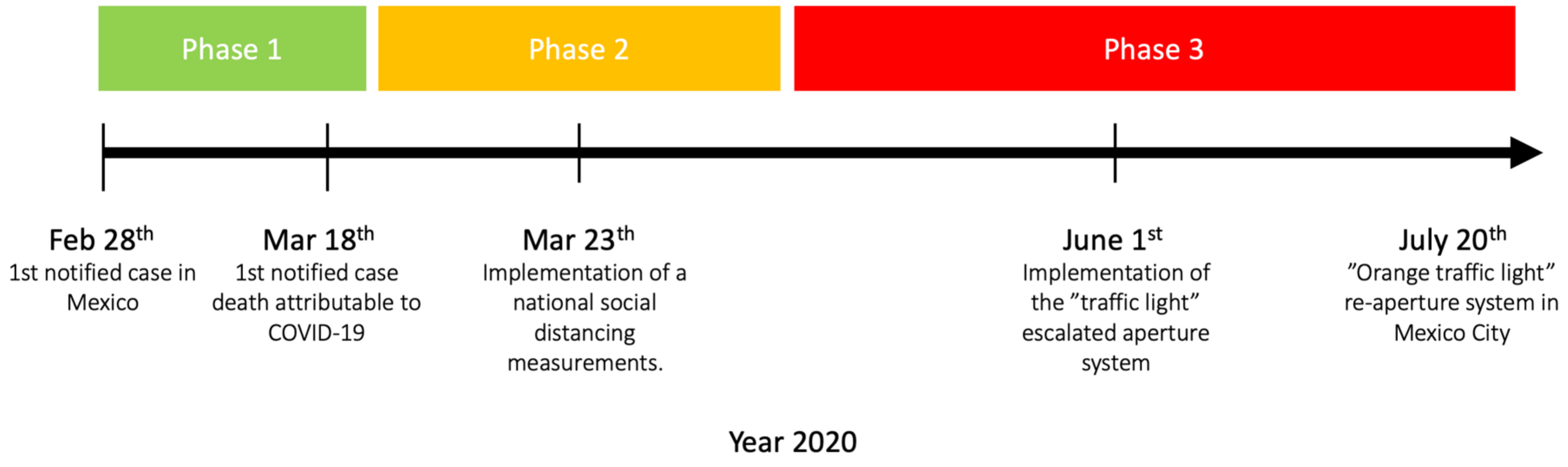

**Supplementary Figure 3:** Positivity rates for SARS-CoV-2 test (A) and percentage of epidemiological surveillance case definition (B) across the evaluated period in Mexico City. Abbreviations: ILI= Influenza-like illness; SARI= severe acute respiratory syndrome.

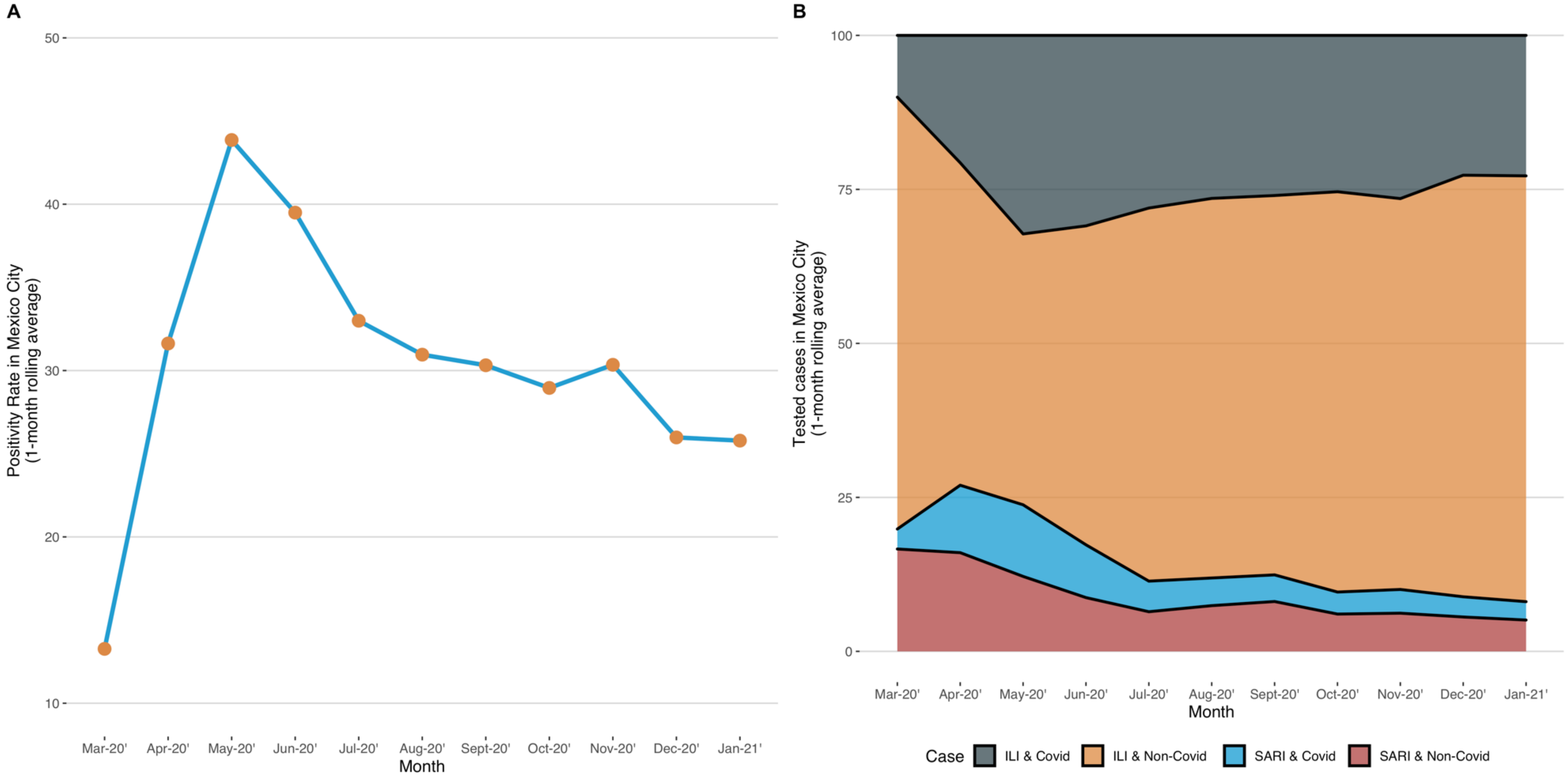

**Supplementary Figure 4:** Population-based analysis of incidence, testing, asymptomatic and mortality rates per 100,000 inhabitants in subjects tested for SARS-CoV-2 infection living in Mexico City.

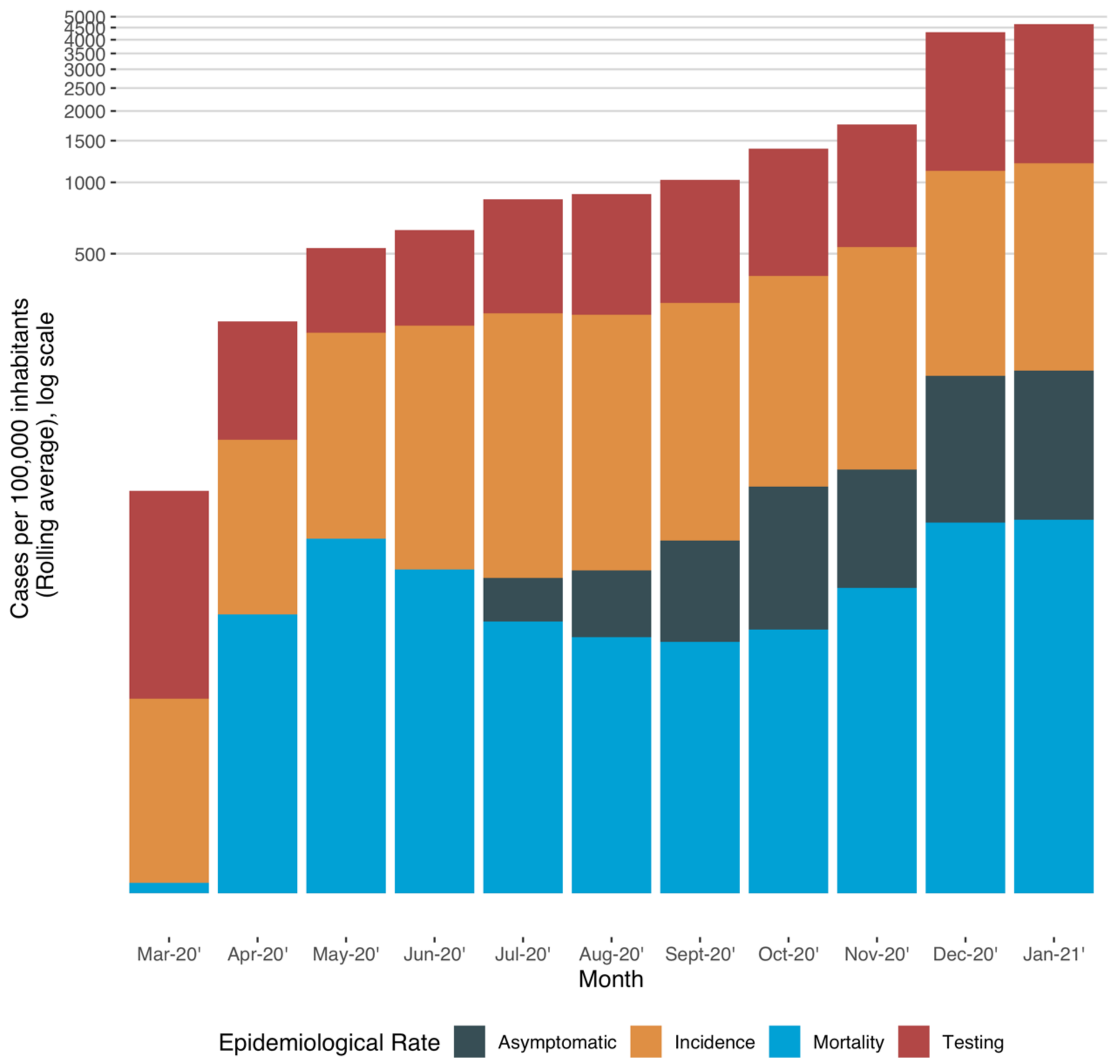

**Supplementary Figure 5:** Positivity rates for SARS-CoV-2 test in working-groups (A) and by SLI-terciles categories (C) with percentage of epidemiological surveillance case definition (B;D) across the evaluated period in Mexico City. Abbreviations: ILI= Influenza-like illness; SARI= severe acute respiratory syndrome.

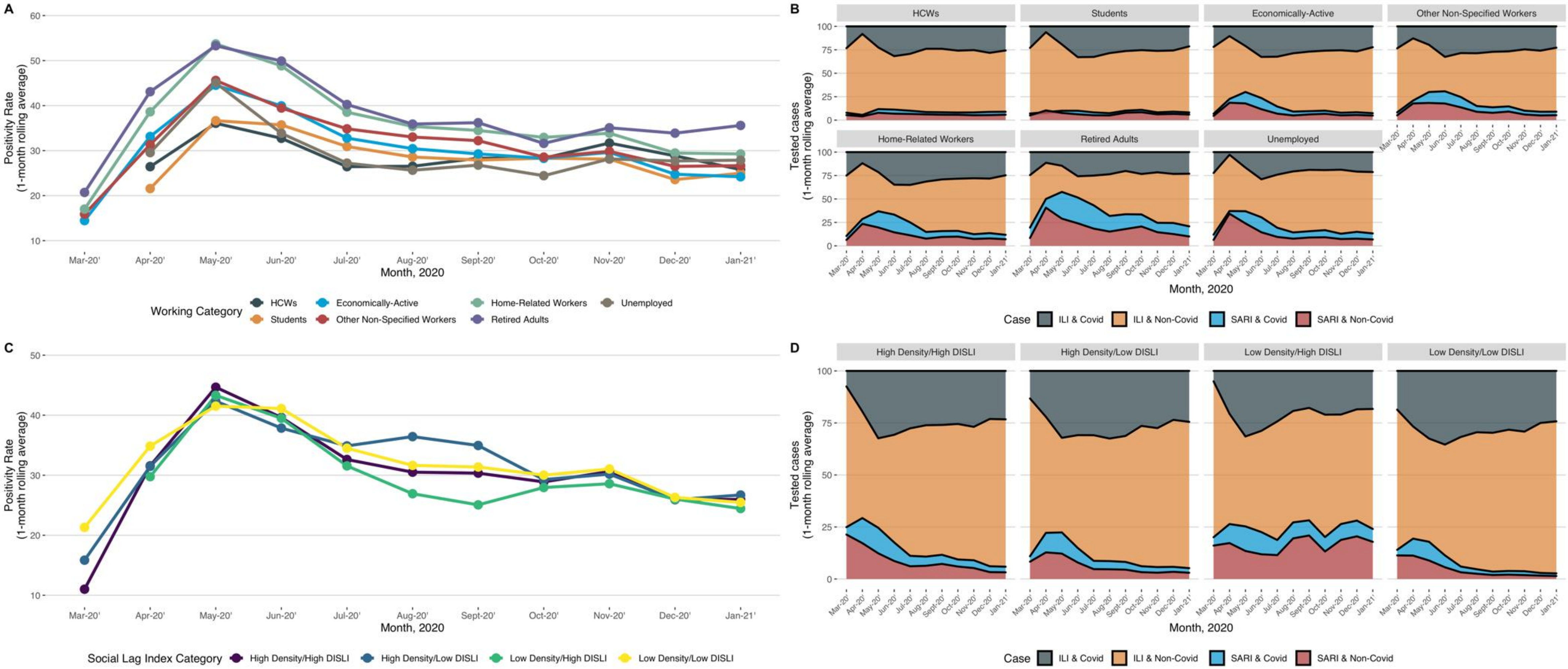

**Supplementary Figure 6:** Analysis of age (A,D,G), number of symptoms (B,E,H) and comorbidities (C,F,I) among the lethal cases stratified by groups of workers. Abbreviations= HIV/AIDS= Human immunodeficiency virus and/or acquired immunodeficiency syndrome; CKD= Chronic kidney disease; CVD= Cardiovascular disease; COPD= Chronic Obstructive Pulmonary Disease.

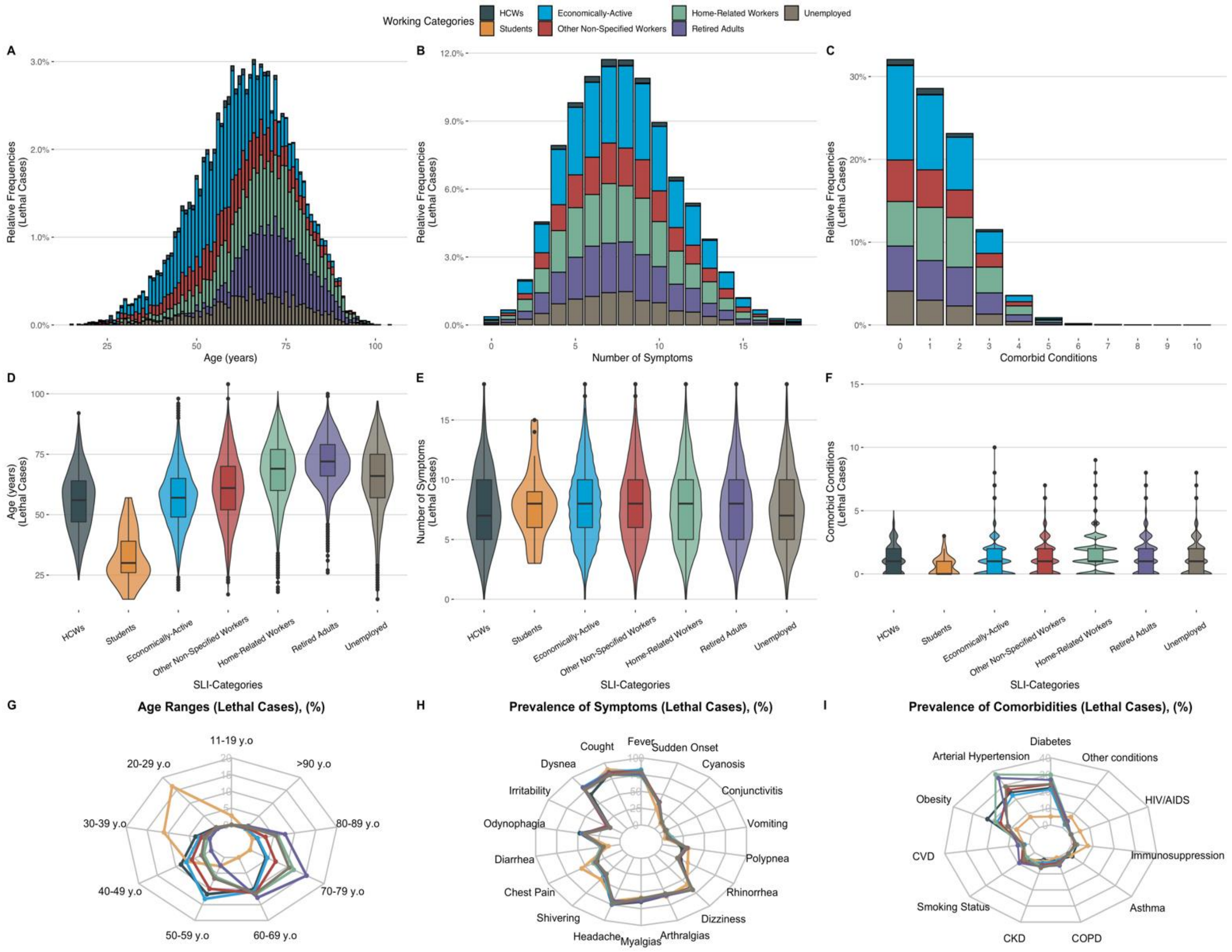

**Supplementary Figure 7:** Analysis of age (A,D,G), number of symptoms (B,E,H) and comorbidities (C,F,I) among the lethal cases stratified by SLI-terciles categories. Abbreviations= HIV/AIDS= Human immunodeficiency virus and/or acquired immunodeficiency syndrome; CKD= Chronic kidney disease; CVD= Cardiovascular disease; COPD= Chronic Obstructive Pulmonary Disease.

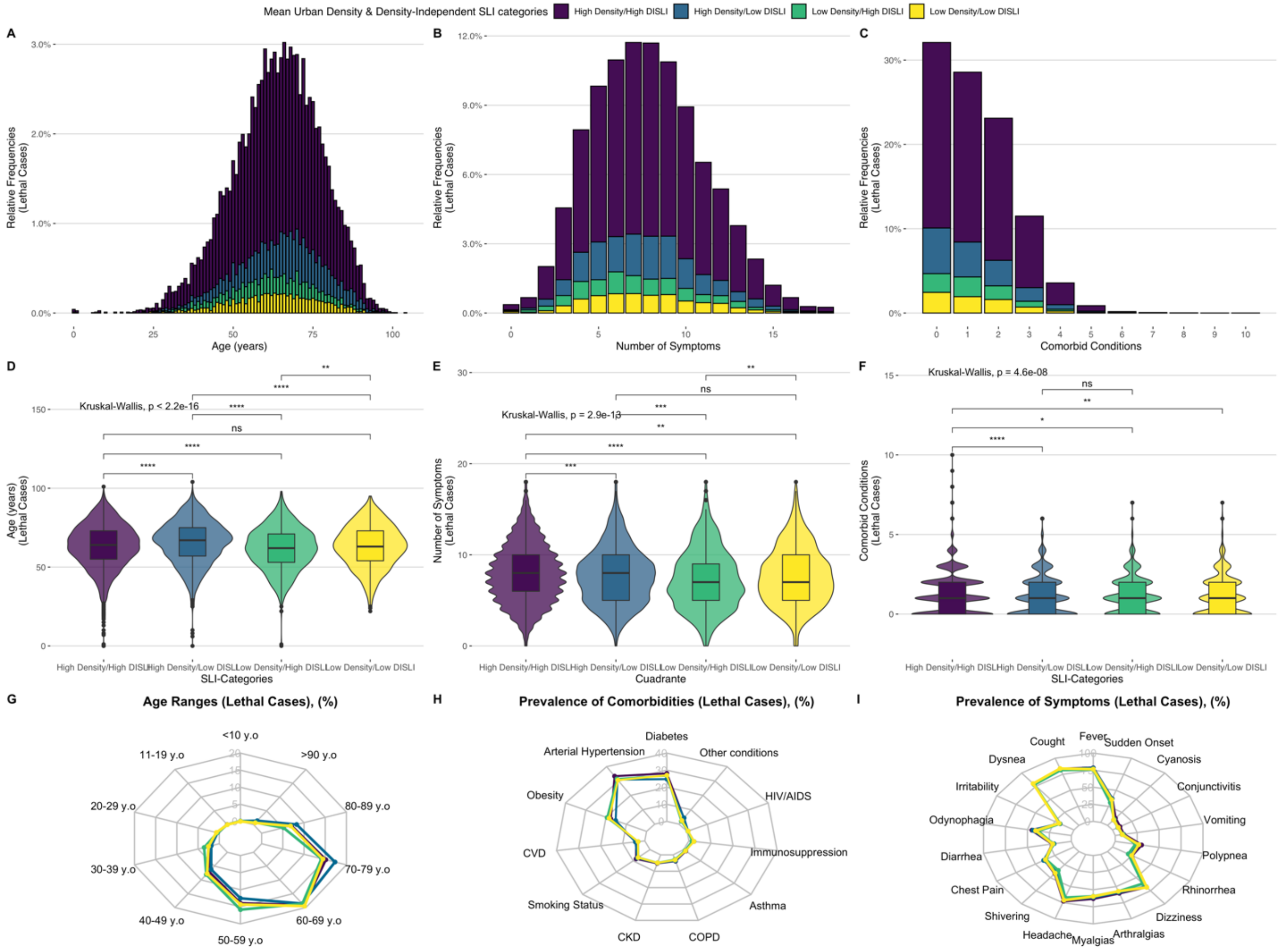

**Supplementary Figure 8:** Interaction effect between working groups treated in ambulatory setting and their risk for lethality attributable SARS-CoV-2 in Mexico City. Abbreviations: SARS-CoV-2 = Severe Acute Respiratory Coronavirus 2

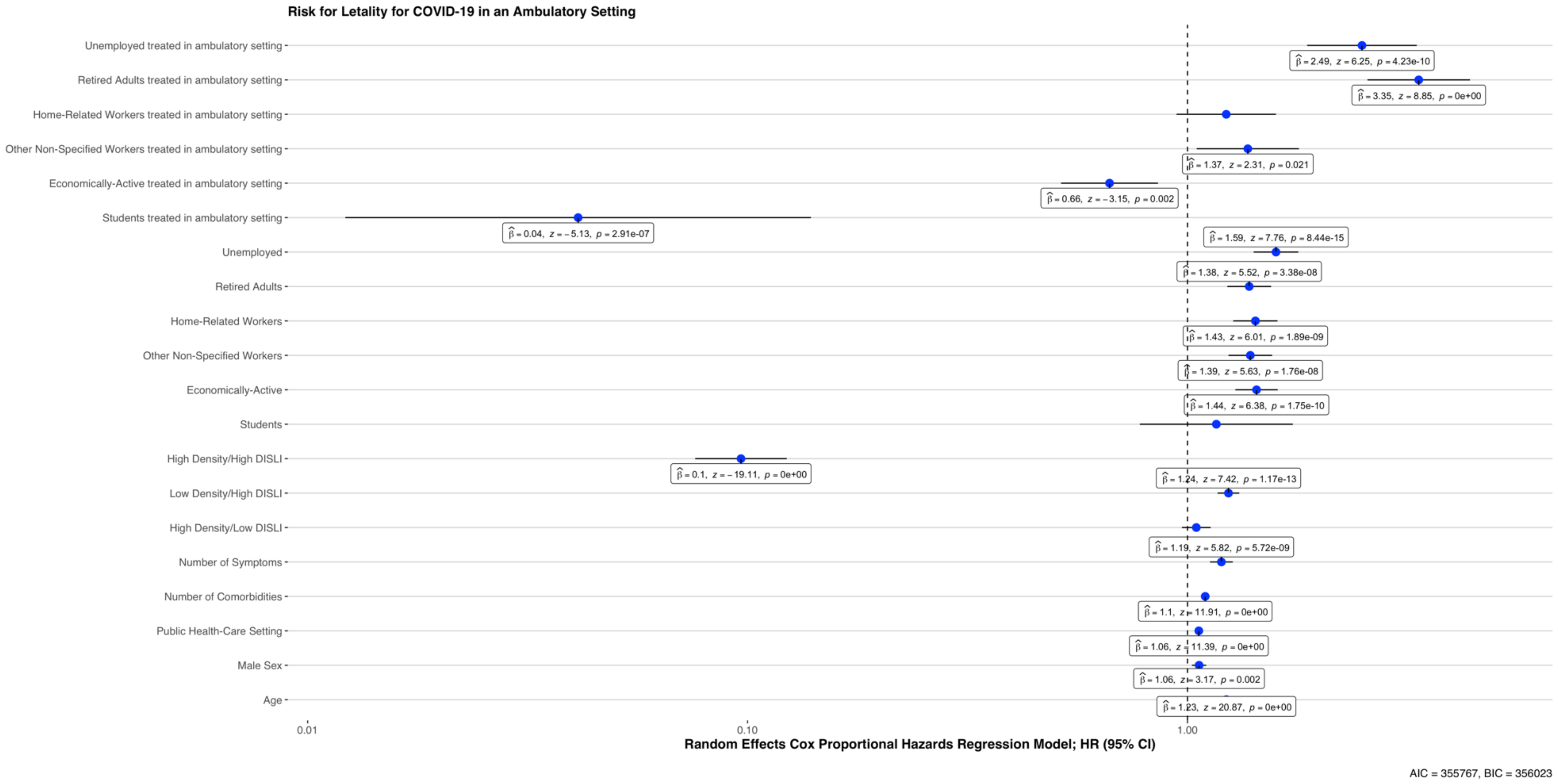

**Supplementary Figure 9:** Relative percent of mobility diminish by municipalities in Mexico City.

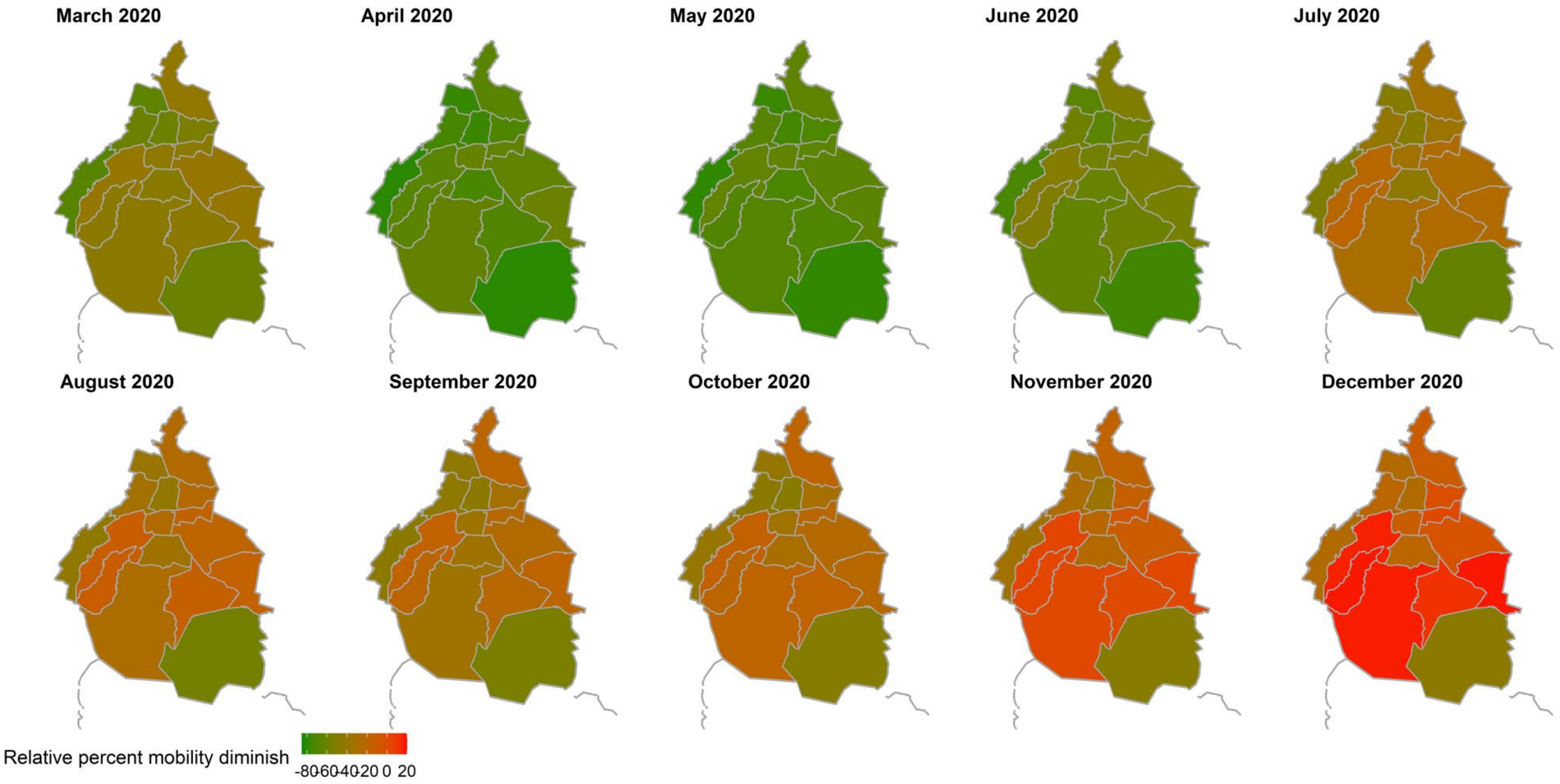

**Supplementary Figure 10:** Poisson regression models to evaluate the interaction effect of vehicular mobility and incidence (A), mortality (B), severe case (C) and hospitalization rate (D) in suspected COVID-19 cases as 31th of December of 2020 by SLI categories in Mexico City..

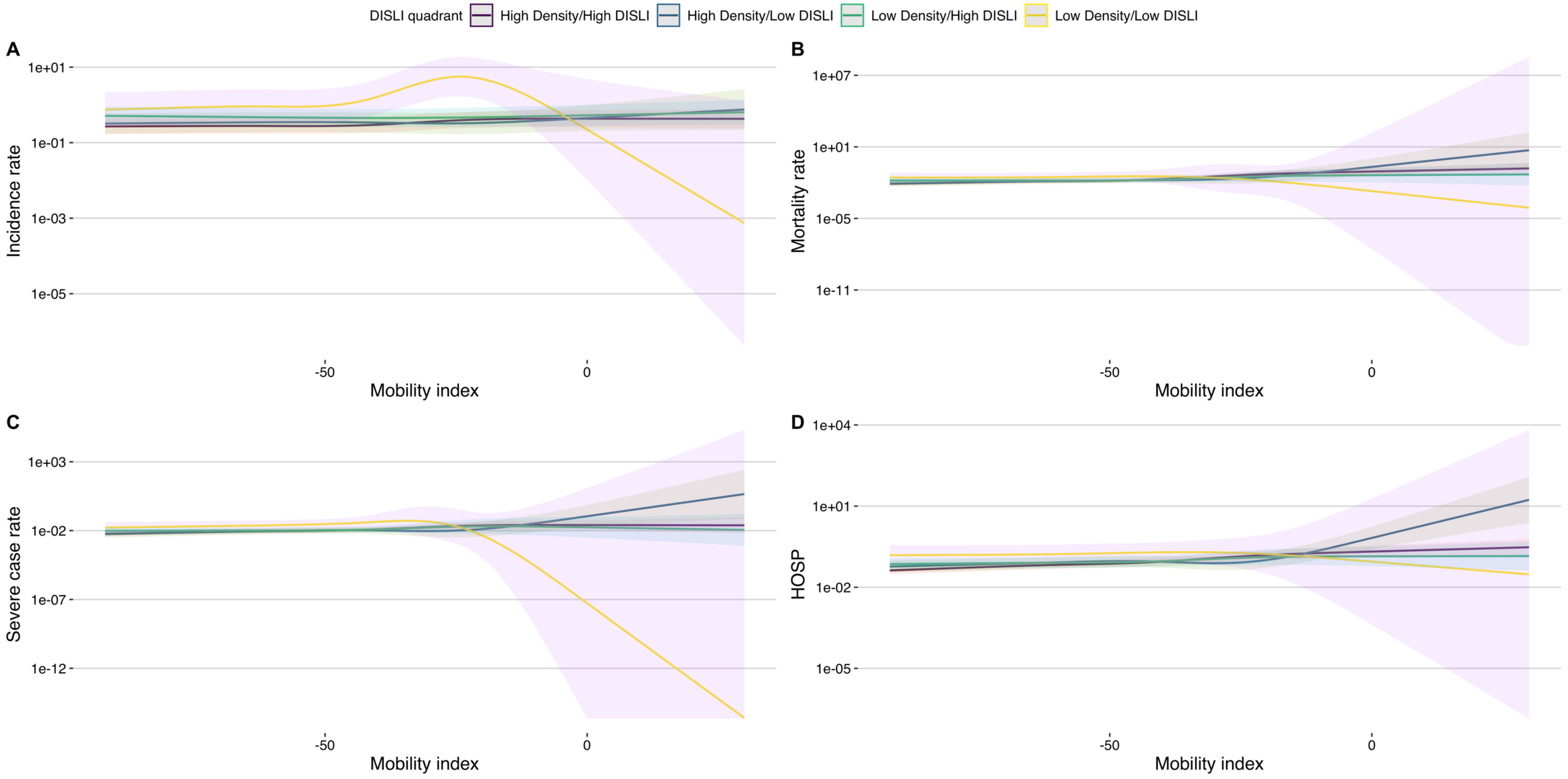

**Supplementary Figure 11:** Age adjusted incidence (A), testing (B), asymptomatic (C) and mortality rate (D) according to mean urban density and density independent social lag index categories in Mexico City.

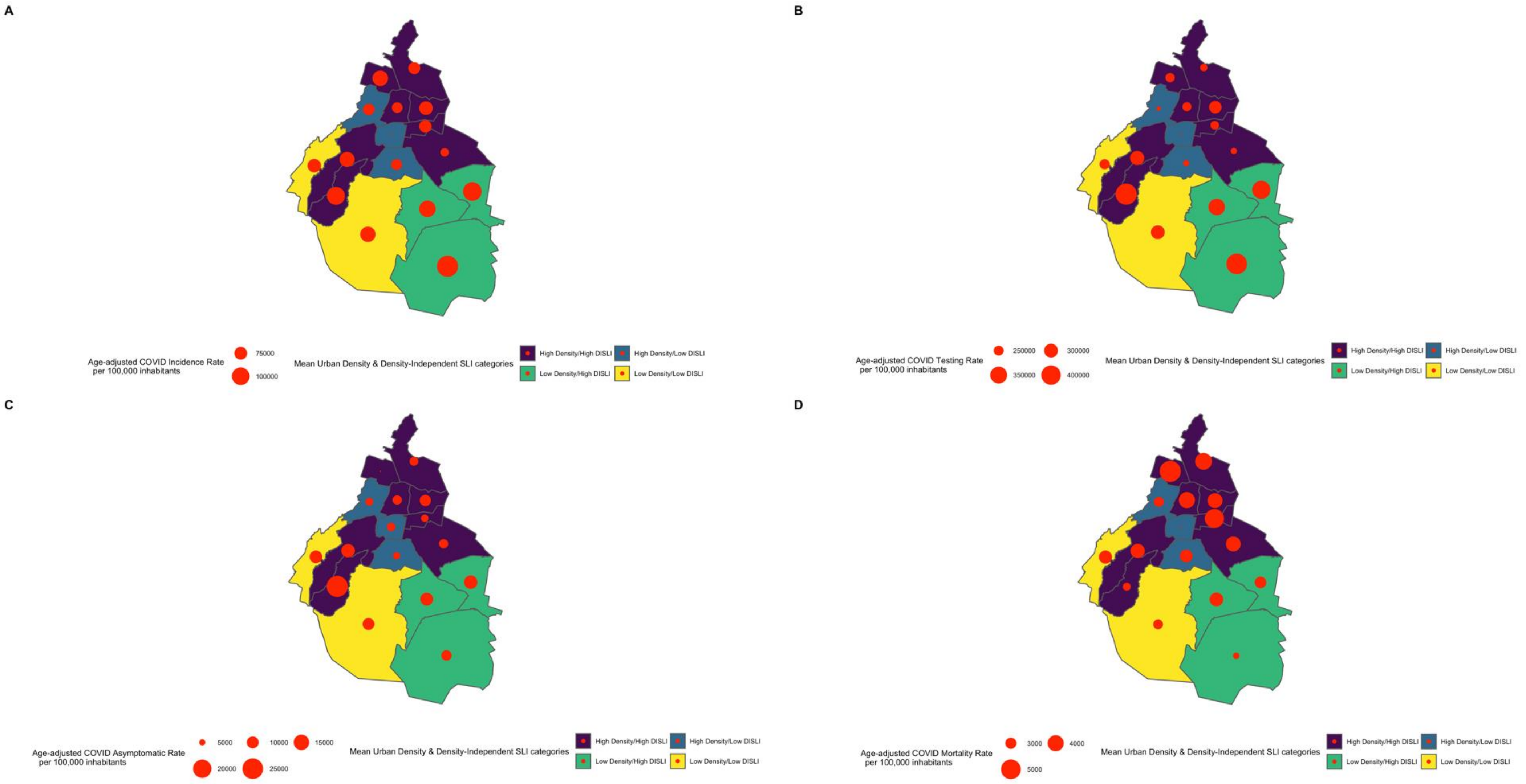

**Supplementary Figure 12:** Age adjusted COVID-19 mortality extracted in death certificates in Mexico City stratified by mean urban density & density independent social lag index categories. Abbreviations: SLI= Social Lag Index

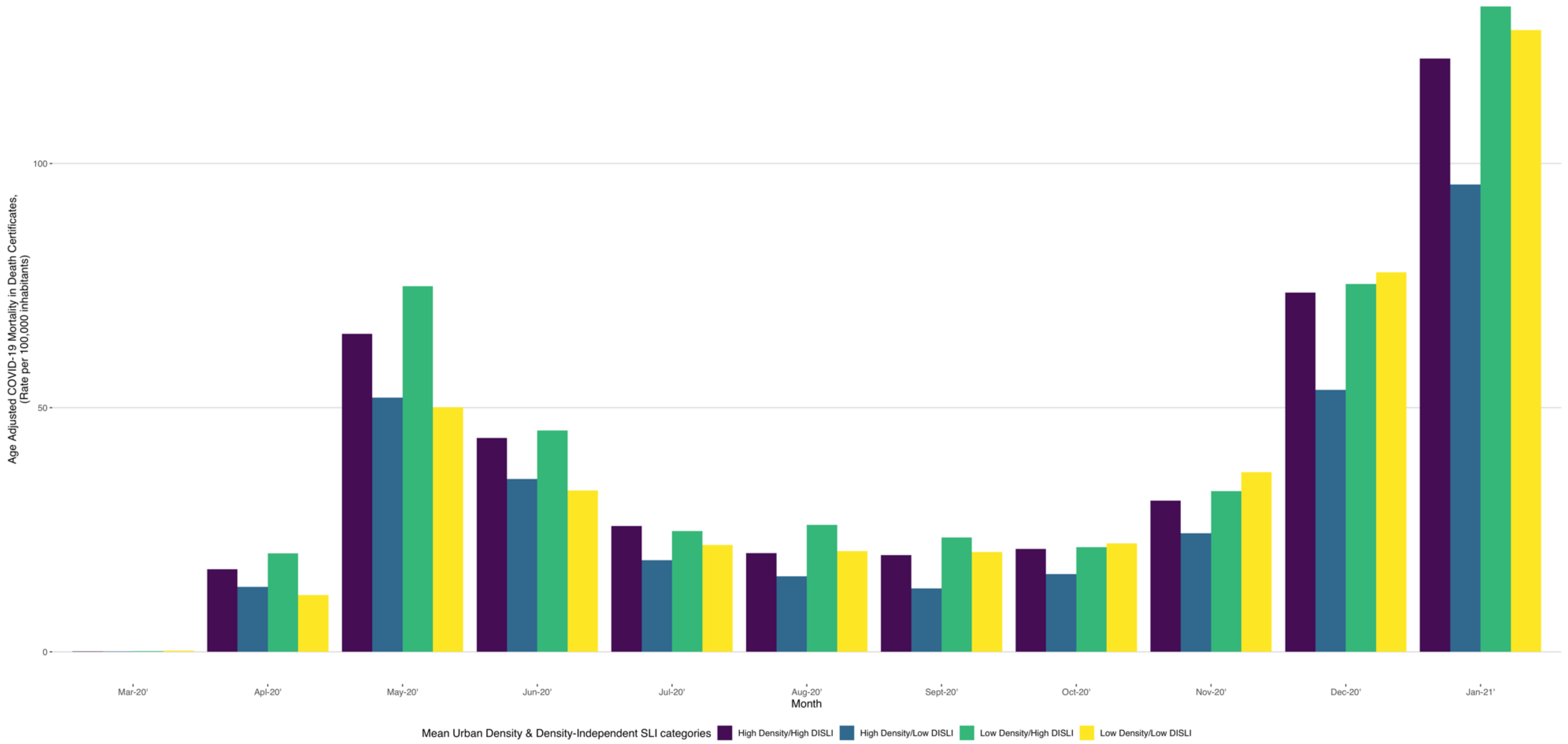

**Supplementary Figure 13:** Rate of each ambulatory death per each hospital non-COVID-19 death by SLI categories (A) and rate of each ambulatory death per each hospital COVID-19 death (B) by SLI categories *Abbreviation*: SLI= Social lag index

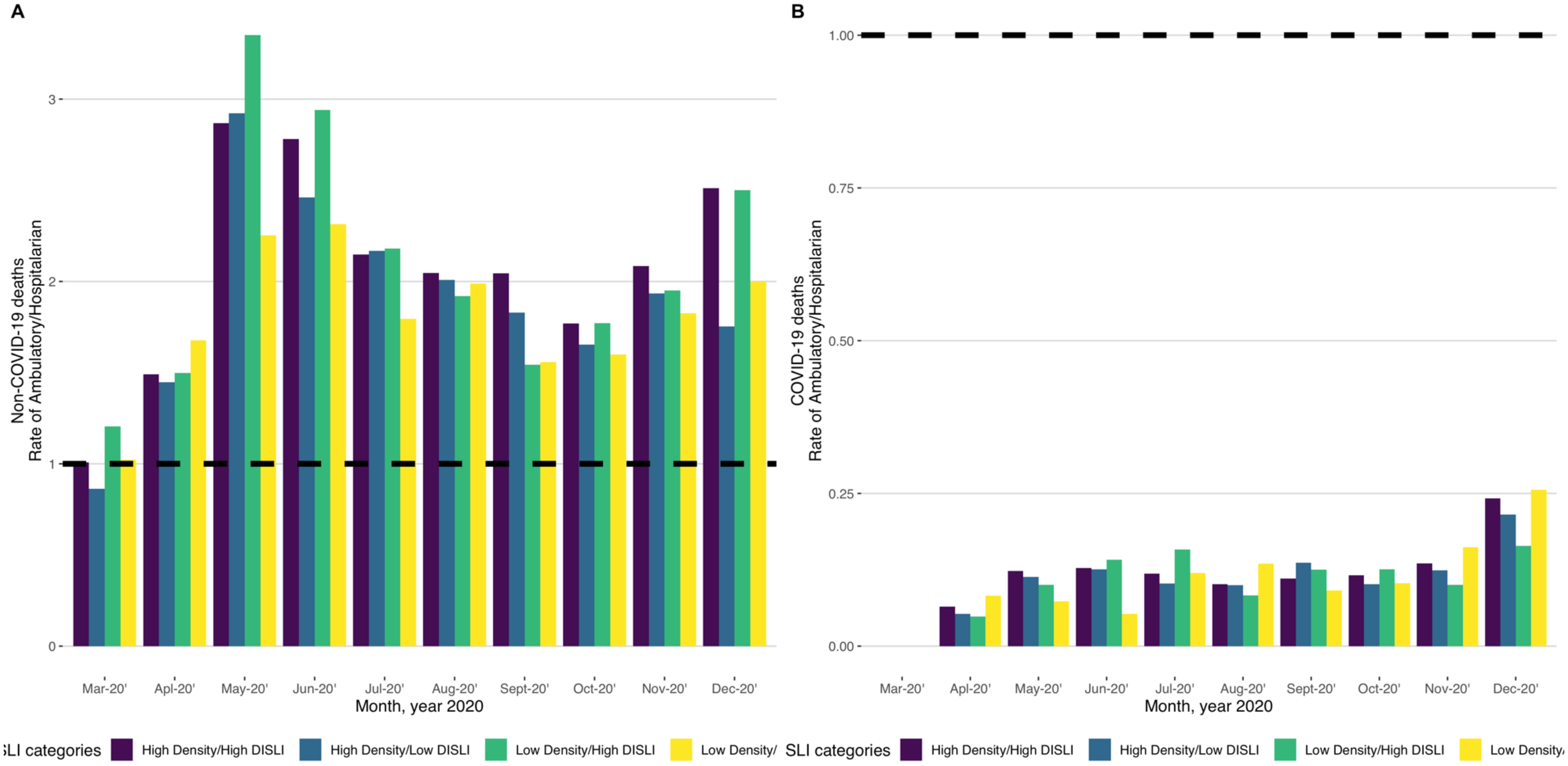

**Supplementary Figure 14:** Non-lineal association of the urban population density independent social lag index (DISLI) with ambulatory-to hospital ratio of COVID-19 deaths (A), non-COVID-19 deaths (B) and with excess mortality rate of non-COVID-19 deaths. *Abbreviation*: SLI= Social lag index

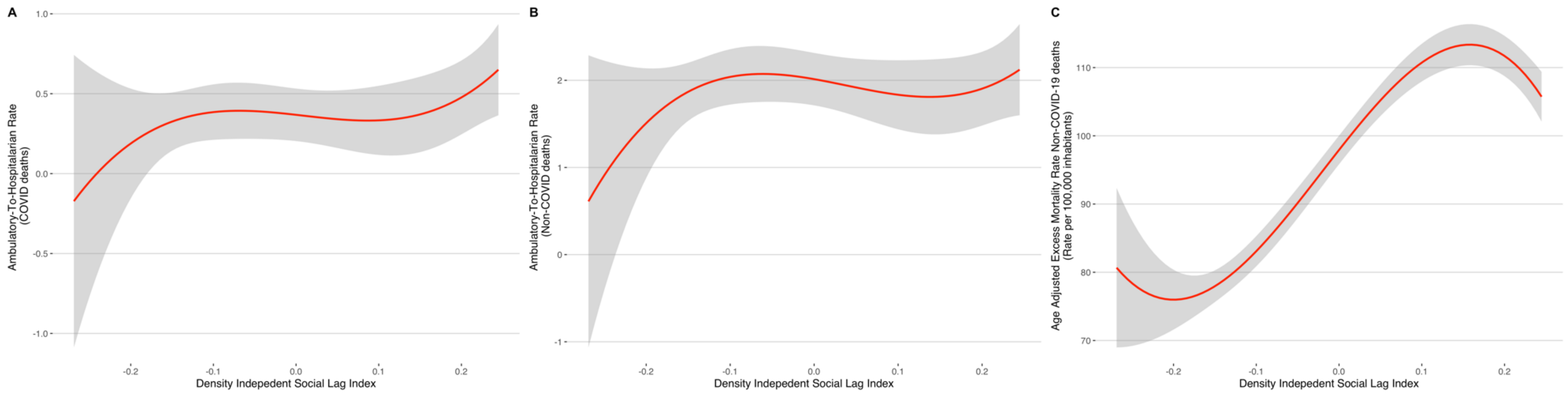
